# Metagenomics disentangles epidemiological and microbial ecological associations between community antibiotic use and antibiotic resistance indicators measured in sewage

**DOI:** 10.1101/2024.12.11.24318846

**Authors:** Connor L. Brown, Monjura Afrin Rumi, Lauren McDaniel, Ayella Maile-Moskowitz, Justin Sein, Loc Nguyen, Minyoung Choi, Fadi Hindi, James Mullet, Muhit Emon, Nazifa Ahmad Moumi, Matthew F. Blair, Benjamin C. Davis, Jayashmina Rao, Anthony Baffoe-Bonnie, Peter Vikesland, Amy Pruden, Liqing Zhang

## Abstract

Wastewater-based surveillance (WBS) is proving to be a valuable source of information regarding pathogens circulating in the community, but complex microbial ecological processes that underlie antibiotic resistance (AR) complicate the prospect of extending WBS for AR monitoring. The epidemiological significance of observed relative abundances of antibiotic resistance genes (ARGs) in sewage is unclear, in part due to multiple sources and in-sewer processes that shape the ARG signal at the entry to the wastewater treatment plant (WWTP). Differentiating between human-derived signals of resistance and those associated with downstream physical and ecological processes could help amplify public health value of WBS of AR by removing noise. In particular, autochthonous sewage microbiota—microbes stably associated with sewage collection networks independent of human/fecal input—could influence profiles of antibiotic resistance via seasonality, temperature, or other factors that alter human community-level AR signals at a given time point. Here we address this fundamental challenge by differentiating distinct associations between sewage-borne antibiotic resistant bacteria and outpatient antibiotic use in the community served by the sewershed. This was made possible using a unique dataset of outpatient antibiotic prescription rates encompassing the majority of antibiotic use over a 5-year period. Leveraging a yearlong 2× weekly sampling of a conventional WWTP with deep metagenomic sequencing (average 29 Gbp/sample) and extensive bioinformatics analysis, we identify striking associations between sewage-borne ARGs and antibiotic usage depending on the putative bacterial host and the presumed environmental stability of the antibiotic. It was found that a subset of ARGs, predominantly associated with *Enterobacteriaceae*, displayed a direct correlation with antibiotic usage, while ARGs predominantly associated with *Pseudomonadaceae* displayed a lagged relationship with antibiotic usage (between 1-3 months). Nested statistical modeling was applied to model the relationship between *Pseudomonas* metagenome assembled genomes and lagged sulfamethoxazole/trimethoprim use while jointly considering sewage characteristics and seasonality. This effort demonstrates the utility of WBS for understanding epidemiological dimensions of AR and provides a framework for accomplishing this purpose by considering microbial ecological factors that contribute to the corresponding signals in sewage.

## INTRODUCTION

Wastewater-based surveillance (WBS) of antibiotic resistance (AR) shows promise in addressing limitations of clinical testing by focusing efforts on composite samples that can capture a multitude of community-level health information (1–3). However, WBS of AR is a fundamentally more daunting task than WBS of individual disease agents, such as SARS-CoV-2. Antibiotic resistant pathogens (ARPs) are numerous and vary widely in their ecology, resistance mechanisms, and epidemiology (4, 5). A general challenge with any WBS effort is that human-associated microbes make up only a small fraction of the diverse microbiome encountered in wastewater (6, 7). The microbiome of municipal sewage is a complex and semi-transient mixture of microbes associated with human excreta, greywater (including microbes washed from the skin), surface and groundwater (infiltration and intrusion), and sewer-associated biofilms and sediment. (6) The fraction of the microbiome not attributable to fecal-material has been found to display strong seasonality associated with shifting wastewater temperature (7). In addition, the unique microenvironment of sewer sediments and biofilms can serve as a reservoir for ARPs, antibiotic resistance genes (ARGs), antibiotics, and other agents that can impose selective pressure (8–10). Thus, sloughed off biofilm and microbiota from sediment most certainly contribute to the AR signal encountered in the influent of a wastewater treatment plant (WWTP). Towards establishing measures of AR in the human community served by the WWTP, a greater understanding of the fecal– and non-fecal sources of AR could help demonstrate the usefulness of WBS in yielding – relevant information and guide formulation of monitoring strategies.

Antibiotic usage and the development and excretion of corresponding resistance determinants in a population is not generally a 1:1 relationship. Multiple studies suggest only weak relationships between antibiotic use and AR using globally-distributed datasets (11, 12). One interpretation of this has been that the transport of resistant microbes, rather than Darwinian selection, can often be the primary force behind observed elevated levels of AR (11, 13). A recent investigation leveraging a global transect of sewage metagenomes likewise observed that antibiotic use was a poor predictor of ARGs encountered in sewage, defined as drug class abundances (i.e., the sum of individual ARGs belonging to a given drug class normalized to 16S rRNA copies) (13). In this study, the authors singled out ecology (defined by the authors as the totality of socioeconomic, climate, and physical chemical aspects related to the environment and sample in question) as a potential mediator of sewage-associated AR. In addition, we recently described a rare case where both outpatient antibiotic prescription rates and community sewage metagenomes were available where there was likewise a limited degree of correlation between antibiotic use and summed ARG drug class abundances (14).

Prior work has suggested that associations between antibiotic residuals and AR are moderated by pre-existing microbial ecological dynamics (15–18) and the potency of antibiotic-driven selection in the specific environment of interest (19, 20). Predicted no effect concentrations (PNECs) of antibiotics are considerably lower in real-world, mixed community conditions relative to clinical breakpoints (21, 22). Along this line, another key consideration is the environmental concentration, stability, and bioavailability of antibiotics (23–25). Whereas the beta-lactam rings in penicillin antibiotics are easily hydrolysable under environmental conditions, macrolides such as azithromycin remain relatively stable (24, 25). Likewise, the folate biosynthesis pathway antagonists sulfamethoxazole and trimethoprim are commonly detected contaminants in aquatic ecosystems. Indeed, trimethoprim and sulfamethoxazole were listed in the 2022 EU watch list of pharmaceutical contaminants posing significant risk to aquatic environments (26). Macrolide antibiotics were also surveilled until 2019 when they reached the maximum four years of data collection (27). To understand the epidemiological signal in wastewater as well as the role of wastewater in selecting for AR, there is a need for research that can address these multiple facets in a holistic manner. Situations where both highly-resolved longitudinal WBS data are collected in tandem with tracking of antibiotic prescription practices and chemical measurements of antibiotics could help shed light on the selective impact of antibiotic use.

Here, we sought to disentangle disparate sources of AR underlying wastewater-borne AR signals to advance the epidemiological value of WBS efforts. We examined the impact of community antibiotic use on sewage-associated microbiota by integrating sewershed-level antibiotic outpatient prescription rates, chemical measurements of antibiotics, and a highly-resolved longitudinal WBS sampling subject to deep shotgun metagenomic sequencing. We hypothesized that discriminating between microbes likely to be excreted in feces from those intrinsic to the water or sewage microbiome (i.e., not fecal-associated) would reveal distinct impacts of antibiotic usage.

## RESULTS AND DISCUSSION

### A highly resolved longitudinal sampling of influent sewage subjected to deep sequencing

Composite samples of sewage influent were collected from a small conventional WWTP (∼ 3.5 MGD) twice weekly for approximately one year (August 2020 to September 2021) **(Figure 1A/B)** and subjected to deep metagenomic sequencing (averaging 29 gigabasepairs (Gbp) or 197 million reads per sample) to produce a total of 3,248 Gbp of Illumina sequencing data **(Supplementary Data 1)**. The primary data collection period coincided with behavioral changes in the area, such as school closures and a shift to online instruction among other COVID-19 interventions **(Fig. 1)**. Differences between community WWTP influent sewage, which is the primary subject of the current study, and the university sewage of a neighboring town were highlighted by changes in the relative abundance of crAssphage, a human fecal marker used as an estimate of the total number of individuals contributing fecal matter to the sewage **(Fig. 1A).** A decrease in crAssphage was observed in November to January as university classes shifted to online only and the semester ended. The sequencing depth achieved here produced substantial coverage (Nonpareil estimated coverage mean 0.74±0.05 with a range of 0.59 – 0.90) **(Fig. 1B).**

**Figure 1.**
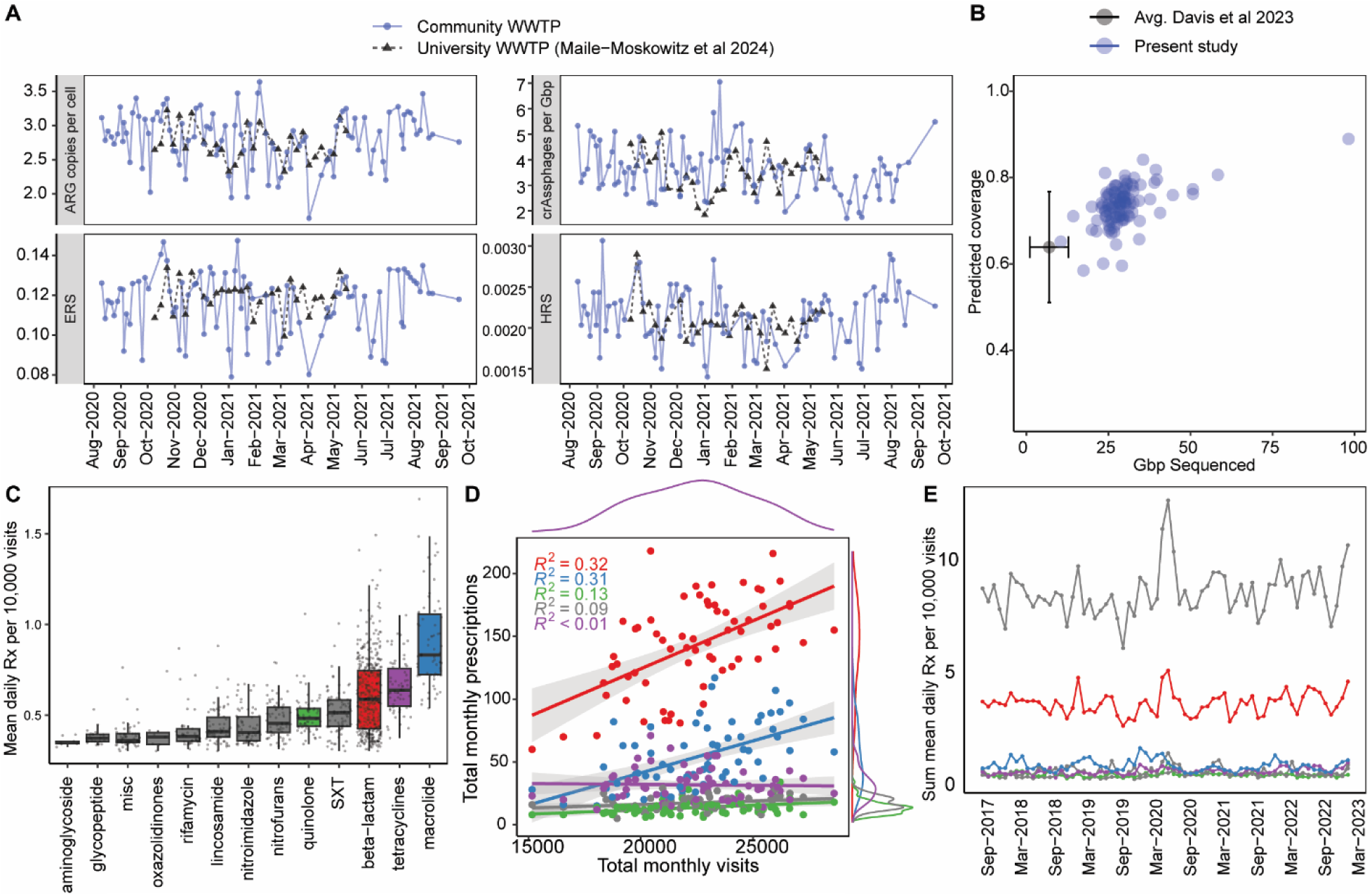
Overview of data included in this study. (A) Characteristics of sewage metagenomes, including total ARG copies per cell (*rpoB* normalized); crAssphage genome copies per Gbp library; MetaCompare 2.0 (28) ecological resistome risk score (ERS); and MetaCompare 2.0 human health resistome risk score (HHRS). Data from the present study are compared to groupings previously established for a neighboring WWTP by Maile-Moskowitz *et al.* 2024. (B) Nonpareil analysis illustrates the sequencing depth of metagenomic data collected for the present study. For comparison, the average library size and predicted coverage for 393 influent metagenomes extracted from the majority of publicly-available data (Davis *et al.* 2023) (29) is plotted. (C) Mean daily rates of outpatient prescriptions for different antibiotics in the sewershed (determined by patient home address, deidentified by zip code). Individual points are daily prescriptions per 10,000 monthly visits (averaged by month). Boxplot summary statistics are: center line: median; upper/lower hinges: 75^th^ and 25^th^ percentiles, respectively; upper and lower whiskers represent the data points extending from the hinge to at most 1.5 times the interquartile range. SXT: sulfamethoxazole and trimethoprim combination therapy. (D) Distributions of prescription rates and visits to outpatient clinics. Each point is the sum of prescriptions for all sub classes within each major category (e.g., beta-lactam). (E) Rates of outpatient antibiotic prescriptions over time (2017–2023). C-E: Includes all time points from 2017-2023.

### Seasonal patterns of antibiotic prescriptions

As was reported previously, antibiotic usage during and directly prior to the wastewater sampling period (Aug 2020 – Aug 2021) was characterized by periodic disruptions due to the COVID-19 pandemic and associated interventions (14) **(Fig. 1)**. Usage data covered 48 outpatient facilities from 2017-2023 and spanned 13 major classes of antibiotics and 27 sub classes. The most prescribed major classes of antibiotics were beta-lactams (especially aminopenicillins and 1^st^ generation cephalosporins), macrolides, tetracyclines, and fluoroquinolones **(Fig. 1C/D)**. Interestingly, there was a spike in prescriptions in March 2020, corresponding with the March 31^st^ pandemic declaration by the World Health Organization **(Fig. 1E).** A total of 19,664 ICD diagnosis codes were associated with antibiotic prescriptions. The most common were acute sinusitis, n=3653 (18.6%); acute bronchitis, n=1159 (5.9%); suppurative and unspecified otitis media, n=1088 (5.5%); acute pharyngitis, n=1055 (5.4%); and cystitis, n=1044 (5.3%) **(Supplementary Data 2)**.

We modeled seasonal patterns of antibiotic usage using the framework of Sun *et al* (30). The model included a sinusoidal component and a linear component that adjusted for year (see methods, **Model 1**). A seasonal pattern was found (amplitude FDR < 5%) for eight of thirteen drug classes (**Supplementary** Fig. 1**, Supplementary Table 1).** These included 1^st^ generation 2-methyl-5-nitroimidazoles, 1^st^ and 3^rd^ generation cephalosporins, aminopenicillins, tetracyclines, and macrolides. Nitroimidazoles, macrolides, and aminopenicillins were associated with the winter months (coded by number, e.g., January = 1). Macrolides showed the most prominent seasonal association (phase: 1.3 95% CI: 0.56 – 2, i.e., January/February; amplitude of 0.23, 95% CI: 0.17-0.30, i.e., about a 20% increase over baseline prescription rates). Aminopenicillins showed a strong seasonal association (phase: 2.5, 95% CI: 1.7-3.2; amplitude: 0.12, CI: 0.69-0.185) followed by nitroimidazole (phase: 0.45, 95% CI: –0.37-1.28; amplitude: 0.04; 95% CI: 0.01-0.07). By contrast, nitrofurans (phase: 3.19, 95% CI: 2.24-4.16; amplitude 0.062, 95% CI: 0.01-0.11), 1^st^ generation (5.40, 95% CI: 3.64-7.15; amplitude: 0.06, 95% CI: 0.01-0.1) and 3^rd^ generation (3.21, 95% CI: 2.50-4.0; amplitude: 0.1, 95% CI: 0.04-0.15), and tetracyclines (5.73, 95% CI: 5.22-6.24) were associated with late spring and early summer months (March to May). Notably, folate antagonists (FA) and 2^nd^ generation cephalosporins did not have detectable seasonality, possibly because of their association with urinary tract infections, which are considered sporadic rather than seasonal **(Supplementary Data 2)**.

In the following months coinciding with sampling, overall outpatient prescription rates were depressed relative to previous years **(Fig. 1)**. Seasonality of prescription rates were assessed while varying which years were included, revealing a depression in seasonal usage of macrolides and 1^st^ generation cephalosporins, but an increased usage of nitrofurans and fluoroquinolone antibiotics during the height of the pandemic (2020–2021) **(Figure 1, Supplementary Figure S1 and S2)**.

### Resistance in sewage-associated microbiota reflects a variable lagging association with antibiotic use

We hypothesized that correlation of individual ARGs with antibiotic use could reveal subtle associations that might not be apparent when considering abundances of ARGs as lump sums. The relative abundance of major drug classes were dominated by only a handful of ARGs, which displayed different overarching trends **(Supplementary** Figure 3). To identify potential associations between AR measures in sewage and community antibiotic usage, we carried out correlation analyses. We noted significant correlations between individual ARGs and antibiotic usage, considering up to three months of lag between antibiotic use and resistance. We considered lag between time of prescription (as a proxy for use) and AR measured in sewage because it is currently unknown where and under what circumstances environmental concentrations of antibiotics impose selection pressure for AR during sewage conveyance. A maximum of three months of lag was considered as this was also applied in a previous study examining relationships between seasonal antibiotic use and resistance (30) **(Supplementary** Figure 4). Multiple points along the lifecycle suggest that cumulative effects (e.g., dosage, length of therapy, environmental stability of the antibiotic, chemical properties of the antibiotic affecting fate) could impact the selective pressure exerted by sewage-associated antibiotics **(Supplementary** Figure 5 **and Supplementary Table 1).**

Following FDR correction, a total of 286 ARGs were found to bear statistically significant associations with antibiotic usage, including negative, positive, lagged, and unlagged relationships **(Supplementary** Figures 6-8**, Supplementary Data 3)**. The hosts of the correlated ARGs were predominantly of the families *Enterobacteriaceae, Burkholderiaceae, Rhodocyclaceae, Morxacellaceae, Aeromonodaeceae,* and *Pseudomonadaceae* (determined via analysis of the metagenomic assemblies) (**Supplementary Figs. S8).**

We hypothesized that associations would differ based on the environmental stability of the antibiotic **(Figure 2)**. Tabulated data on antibiotic prescription rates and measurements of antibiotics from non-target pharmaceutical screening analysis **(Table 1)** were used to identify antibiotics with plausible contributions to AR in sewage, on the basis that they were both commonly prescribed and detected in sewage. This analysis highlighted azithromycin, sulfamethoxazole/trimethoprim, and ciprofloxacin as being both commonly prescribed and detected in the sampled wastewaters **(Table 1)**. It should be noted that except for imipenem, beta-lactams were not included in the suspect screening because of their environmental instability (14). In addition, imipenem was not found in the outpatient antibiotic use data due to its intravenous formulation and the fact that meropenem is the preferred carbapenem in the local area.

**Figure 2.**
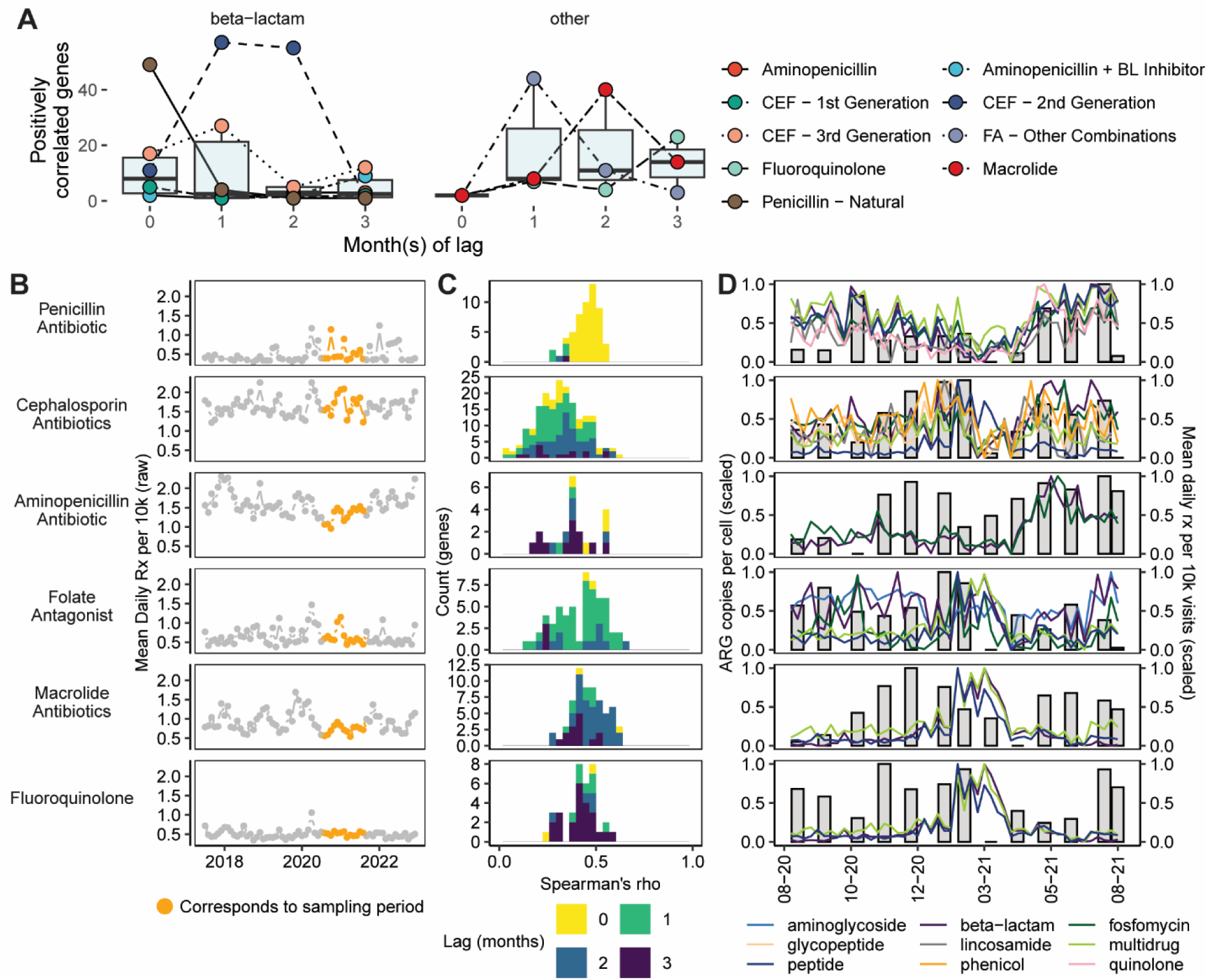
Association of ARGs with antibiotic prescription. (A) Number of genes associated with a given antibiotic partitioned by general and specific drug class of the prescribed antibiotic (not the ARG). The “other” category refers to antibiotics detected in sewage and commonly prescribed in outpatient clinics in the sewershed CEF: cephalosporin antibiotic. BL: beta-lactamase inhibitor. **(Table 1).** Boxplot summary statistics are: center line: median; upper/lower hinges: 75^th^ and 25^th^ percentiles, respectively; upper and lower whiskers represents the data points extending from the hinge to at most 1.5 times the interquartile range. (B) Rates of prescription for each sub class of antibiotic in outpatient settings in the sewershed. Orange regions indicate overlap with the sewage sampling period. (C) Distribution of Spearman’s correlation coefficients for different drug classes and lag. (D) Min-max scaled ARG drug class abundance (summed across all genes; first axis, colored lines) and min-max scaled monthly antibiotic prescription rates (second axis, grey bars).

**Table 1.** Antibiotic prescriptions and corresponding measurements from the pharmaceutical and personal care product (PPCP) screening.

|  |  |  |  |  |  |  |
| --- | --- | --- | --- | --- | --- | --- |
| Drug | n | Days of therapy (DoT, mean ± sd) | Dose Amount (mg, mean ± sd) | Peak Area (measure of relative concentration [mean ± sd]) | Rank (use)* | Rank (peak area) |
| azithromycin | 2850 | 7.83 ± 7.35 | 253.6 ± 57.61 | 179477 ± 265363 | 1 | 3 |
| sulfamethoxazole | 1060 | Dosed twice per day, 12.7 ± 23.7 | 750.2 ± 152.8 | 15012 ± 15488 | 2 | 4 |
| trimethoprim | 1060 | | 100 ± 0 | 1143952 ± 1131570 | 3 | 1 |
| ciprofloxacin | 576 | Dosed twice per day, 7.56 ± 3.96 | 437.9 ± 109.1 | 117513 ± 159465 | 4 | 2 |
| clarithromycin | 198 | Dosed twice per day, 23.5 ± 16.7 | 468.4 ± 88.8 | 36887 ± 85390 | 5 | 5 |
| oral vancomycin | 28 | Dosed four times per day, 13.9 ± 13.0 | 125 ± 0 | 996 ± 1268 | 6 | 6 |
| erythromycin | 12 | Dosed four times per day, 14 ± 7.21 | 380.5 ± 114.6 | 8318 ± 13975 | 7 | 7 |

\*Considers only antibiotics that were also measured in the PPCP screening.

Beta-lactam antibiotics were expected to bear notable direct correlation (i.e., without lag) to corresponding ARGs because of their noted instability in water environments **(Figure 2A-D)**. Indeed, prescriptions for penicillin antibiotics were directly correlated with more than 49 ARGs, but only 6 when usage was lagged **(Figure 2A/C)**. By contrast, macrolides displayed an opposite tendency with respect to lag **(Figure 2A)**. Only two corresponding ARGs were positively correlated with macrolide prescriptions without lag, while 46 were positively correlated at *k* = 2 months of lag **(Figure 2A)**. Similarly, prescriptions of antibacterial folate antagonist combinations were correlated with only two ARGs without lag and 44 ARGs at *k* = 1 month of lag **(Figure 2A).** This result was statistically significant (two-tailed Fisher’s exact test, with groupings of beta-lactam or high environmental relevance drug class; categories corresponded to correlation at *k =* 0 and *k >* 0, while values were the count of genes positively correlated at that lag value; p < 0.0001). Significant overlap in associations between drug classes and ARGs was observed **(Fig. 2D**); for example, macrolide antibiotics and fluoroquinolone antibiotics displayed similar trends with respect to multidrug and fosfomycin ARGs.

One surprising exception to this trend involved 2^nd^ generation cephalosporins, which were almost exclusively prescriptions of cefuroxime **(Figure 2B-D).** Whereas most beta-lactam antibiotics displayed unlagged associations with ARG abundance, resistance was most prominently associated with prescriptions of 2^nd^ generation cephalosporins with two months of lag. It is not possible to know whether the observed trends are truly due to differences in the cephalosporin antibiotics and associated selection pressure. Indeed, cefuroxime was the most scarcely prescribed cephalosporin. Nevertheless, it is noteworthy that cefuroxime has been identified on multiple occasions as a relevant environmental contaminant, including in raw sewage and treated wastewater (31) as well as wastewater biosolids and soil (32–35). In particular, cefuroxime sorbs to a variety of soils in a nearly irreversible fashion (desorption rates of <1%) (32).

Overlaying the putative hosts of significantly associated ARGs onto the results of the correlation analysis revealed a striking pattern in the trajectory of ARGs encoded by *Enterobacteriaceae* and *Pseudomonadaceae* **(Fig. 3a).** This trend was consistent across multiple drug classes and ARG mechanisms **(Figs. 3b, Supplementary** Figures 7 and 8). In particular, it was observed that many ARGs predominantly encoded in *Pseudomonadaceae* were significantly positively correlated with antibiotic usage with increasing lag. By contrast, most *Enterobacteriaceae* ARGs were positively correlated only when there was no lag. However, with increasing lag, this trend was reversed with most *Enterobacteriaceae* ARGs being negatively correlated while *Pseudomonadaceae* became positively correlated most prominently at two months of lag, suggesting a turnover in host responsiveness to antibiotic use **(Fig. 3A)**. Taxonomic separation with respect to lag suggested the potential for effects of antibiotic use and excretion that impacted some wastewater microbiome constituents differently than enteric-associated organisms **(Fig. 3A, Supplementary** Figure 7). We predicted ARG co-abundance groups (CAGs) based on their relative abundance (Methods) **(Fig. 3B)** and the resulting all-versus-all correlations. This was supportive of at least four CAGs that, in part, were correlated with predicted host taxonomy. ARGs associated with *Enterobacteriaceae* and *Pseudomonadaceae* were largely partitioned into separate CAGs while the remaining top ARG hosts fell across multiple CAGs **(Fig. 3C)**.

**Figure 3.**
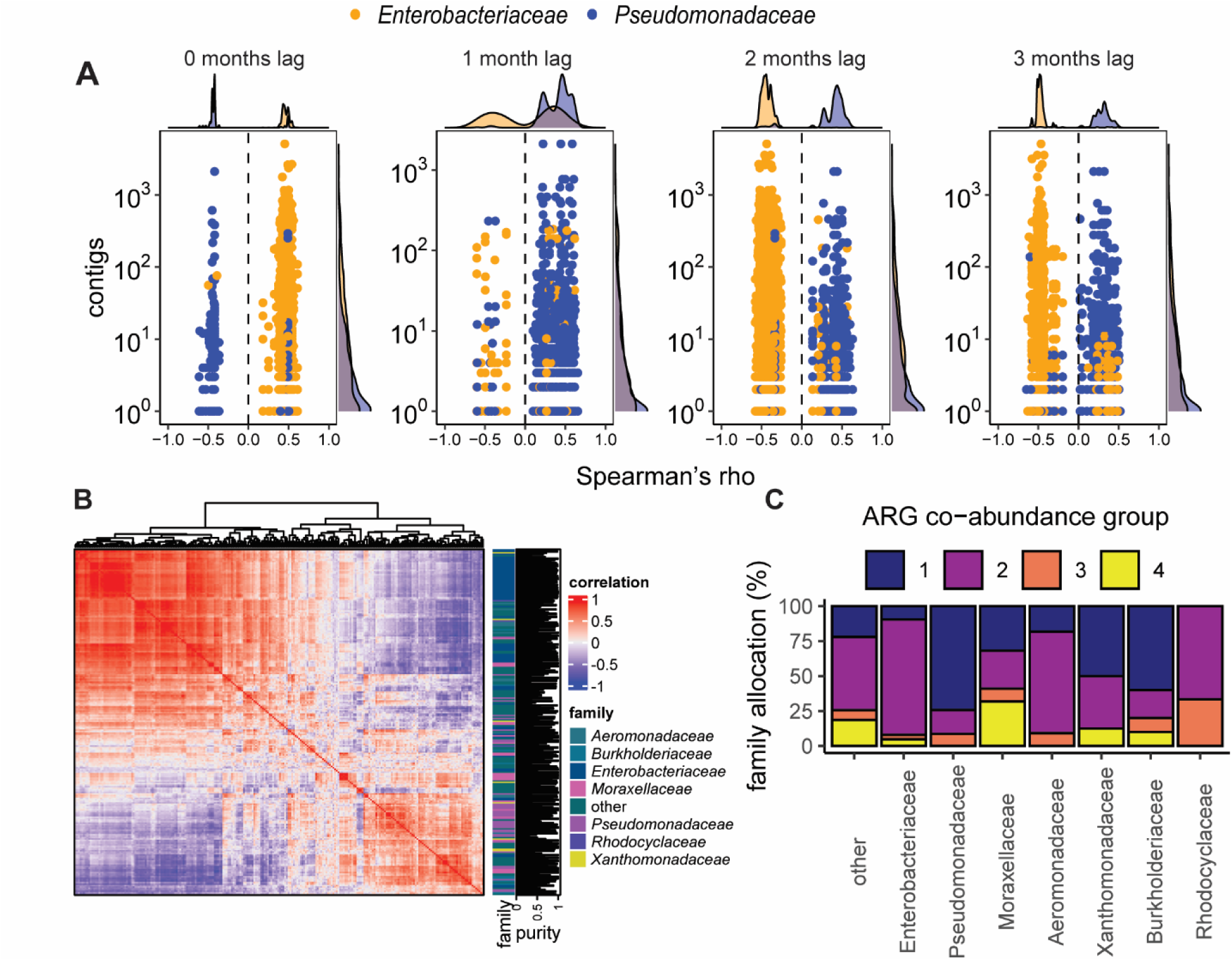
Sewage-associated microbiota have a variable lagging association with antibiotic usage. (A) Significantly correlated ARGs overlaid with host taxonomy (subset to include only *Enterobacteriaceae* and *Pseudomonadaceae*) suggests a turnover in the relationship between usage and the hosts of ARGs over three months. The same figure with the top seven families is available (**Fig. S8**). (B) Heatmap depicting the correlation between ARGs found to have a significant association with antibiotic usage. Each column and row is an ARG, and the heat is the Spearman correlation coefficient between the two genes. Family is the majority-rules family of the ARG and purity refers to the proportion of contigs with a given ARG that are assigned to the depicted family or 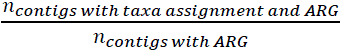. (C) Association of different bacterial families to ARG co-abundance groups. Family allocation is calculated as the proportion of ARGs belonging to a taxon in a co-abundance group to total ARGs belonging to a given taxon.

### Complex population dynamics of ARG hosts in sewage

The pattern observed here (**Fig 3a)** led us to next examine the population dynamics of ARG-bearing genera using the assembly catalogue and the collection of metagenome assembled genomes associated with influent samples (MAGs) (n = 1,890), **Supplementary Data 4**. Unlagged associations were dominated by the order *Enterobacteriaceae,* which includes many (though not exclusively) enteric organisms.

The composition of the sewage microbiome was heavily influenced by seasonality and/or autocorrelation **(Fig. 4).** This was apparent when overlaying the time of sampling **(Fig 4A)** and upon correlating the first MDS axis with temperature **(Fig 4B/C)** (Pearson’s, *R^2^* = 0.72). Interestingly, the second MDS axis correlated well with the abundance of crAssphage (Pearson’s, *R^2^* = 0.51) **(Fig. 4D)** and, similar to crAssphage itself, did not display substantial autocorrelation **(Fig 4D/E, Supplementary** Figure 9). Differentiation of MAGs into co-abundance groups also was suggestive of seasonal turnover in the dominant species of bacteria **(Fig. 4F/G).** Four major co-abundance groups were also observed (Methods) for the MAGs, and, in a similar result as seen for the ARG co-abundance groups, each cluster spanned multiple, overlapping taxonomic groups **(Fig. 4H).** CAG-1 (41 unique genera) displayed peak abundance in summer months and included MAGs classified to one of the ESKAPE-pathogen genera: two *Klebsiella* genomospeices (gs.) and *Pseudomonas* gs.*; Enterobacter* gs.; and *Acinetobacter* gs. CAG-2 was the most diverse cluster (56 unique genera) and displayed peak abundance in winter/spring months and included *Pseudomonas gs.* CAG-3 (29 unique genera) and CAG-4 (20 unique genera) were comparatively stable and, interestingly, CAG-4 included a *Klebsiella* gs. Two genera (*Zooglea* and *Acinetobacter*) were detected across all CAGs **(Supplementary** Figure 10).

**Figure 4.**
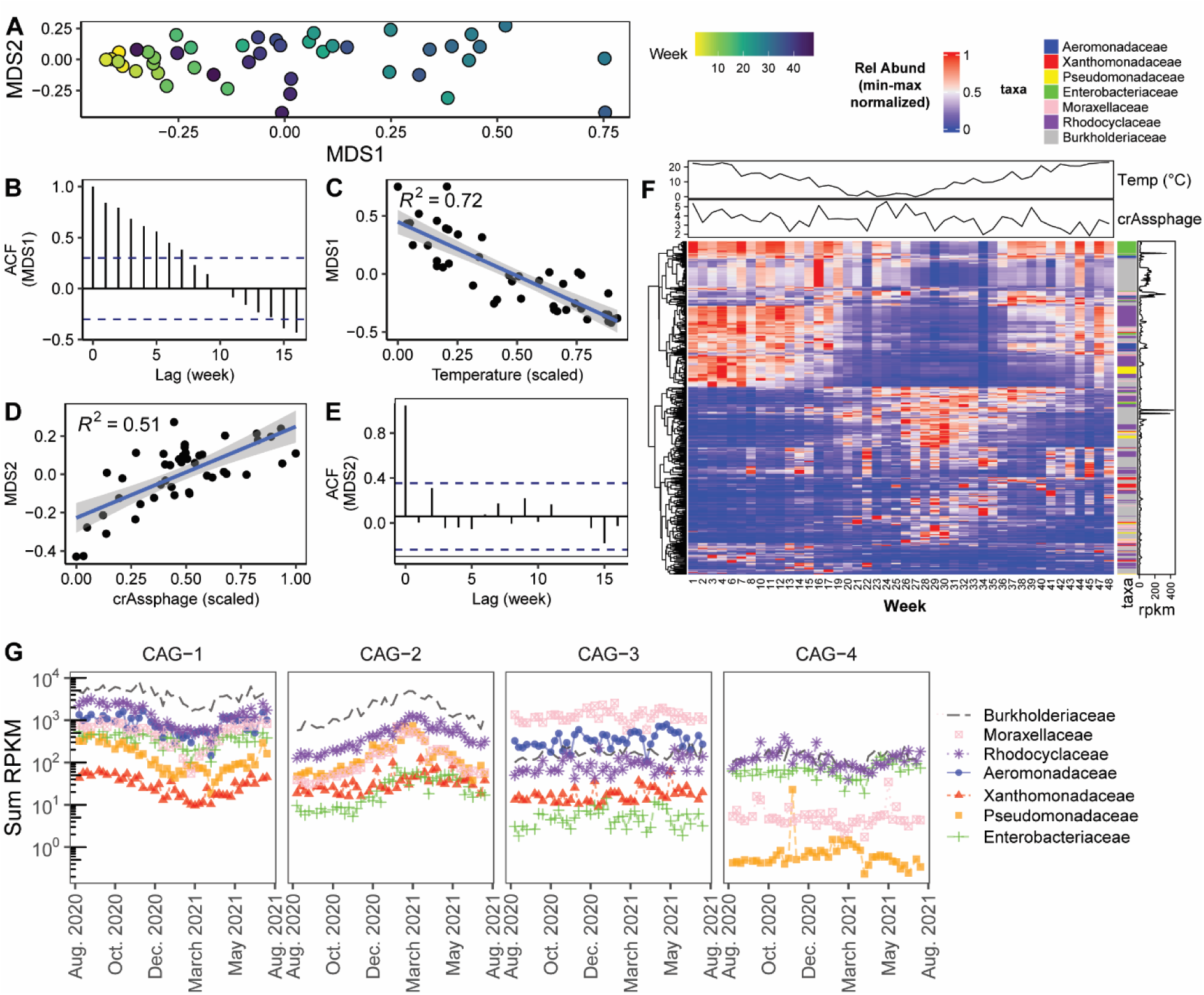
Population dynamics of ARG hosts are related to seasonal fluctuations and the abundance of crAssphage. (A) NMDS ordination of MAG relative abundances for individual weeks. (B) Autocorrelation function (ACF) of the first component (x-axis) in panel 4A. The y-axis relays the correlation of MDS1 with itself by the given value of lag (x-axis). (C) Correlation between min-max scaled temperature and the first component (x-axis) of panel 4A. (D) ACF of the second component (y-axis) in panel 4A. (E) Correlation of min-max scaled crAssphage relative abundance and the second component (y-axis) of panel 4A. (F) Heatmap of minmax normalized MAG abundances by week with row annotations: MAG family and mean RPKM; and column annotations of mean weekly temperature (in °C) and relative abundance of crAssphage in genome copies per Gbp. (G) Summed abundances of all MAGs of the given bacterial family partitioned by co-abundance groups with opposite seasonal trends. (B) and (E): blue lines indicate a 95% confidence interval for white noise data (i.e., timeseries with no autocorrelation).

### Relationship between folate antagonists and *Pseudomonas* spp

Autocorrelation (i.e., non-independence of counts between timepoints) complicates correlation-based analyses (36). For example, AR indicators measured in sewage and antibiotic use might be correlated due to partially overlapping seasonality or the non-stationarity of the data. Simultaneous seasonal shifts in the non-fecal wastewater microbiome and prescription practices could plausibly translate to similarities that are not due to selective pressure. To address this, we performed a nested modeling procedure to jointly assess the impact of antibiotic use and factors associated with seasonality using *Pseudomonas* spp. and folate antagonists **(Models 2/3)**. We argue that jointly modeling periodicity of specific ARG hosts and antibiotic usage reduces the potential for misleading results.

We focused on the relationship between *Pseudomonas* species and folate antagonists because the most common prescription was a combination of sulfamethoxazole (detected in influent from the chemical suspect screening) and trimethoprim (the most abundant antibiotic in the chemical suspect screening) **(Table 2)** and the striking lagged association with usage **(Fig. 2A)**. Sulfamethoxazole and trimethoprim combination therapy (SXT) targets two steps in folate biosynthesis, an essential step in purine metabolism, disrupting cell growth (37). *Pseudomonas* spp. are expected to have a degree of intrinsic resistance owing both to membrane impermeability and chromosomally-encoded efflux pumps (38). However, susceptibility to trimethoprim in *Pseudomonas aeruginosa* has been documented under certain conditions (39, 40). We also detected multiple contigs putatively labeled as *Pseudomonadaceae* with genes aligning to *dfrA* family members from the CARD database **(Supplementary Data 5).** Further, one study previously found correlations between *dfrB* family genes and taxa of the order *Pseudomonadales* in surface waters and wastewater (41). In this case, the presumptive ecological conditions being modeled are the simultaneous occurrence of seasonal turnover in the dominant strains of *Pseudomonas* and selection for FA resistance.

A total of 43 species-level dereplicated *Pseudomonas* gs. MAGs spanning 11 different species, including *P. fluorescens, mohnii, peli, coleopterorum* and 19 MAGs without species-level assignments were recovered **(Figure 5)**. Seasonal-based modeling was deemed appropriate for all species except *P. coleopterorum* **(Figure 5)** based on trajectories in relative abundance **(Figure 5).** Because no substantial variation was observed within *P. alcaligenes* MAGs **(Supplementary** Figure 11), we summed up the relative abundance of individual *P. alcaligenes* gs. to fit one model encompassing all *P. alcaligenes*. We also modeled the abundance of specific MAGs in response to antibiotic use while adjusting for additional explanatory variables **(Model 4)**, such as crAssphage, influent flow, and whether the sample was collected on a Monday. Samples collected on Mondays displayed a moderately elevated resistome relative to samples collected on Fridays **(Supplementary** Figure 12).

**Figure 5.**
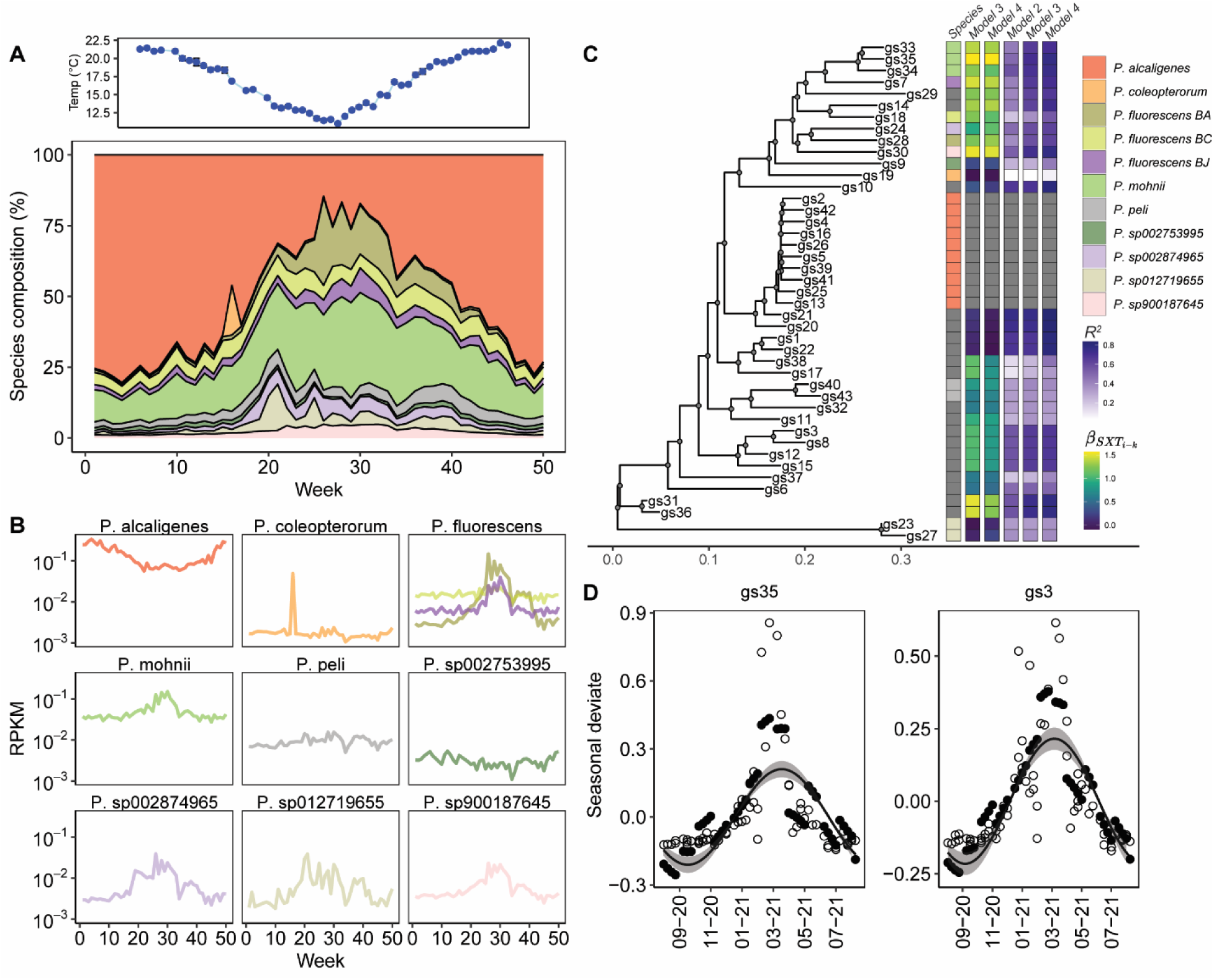
Relationship between *Pseudomonas* genomospecies, seasonality, and antibiotic use. (A) Average weekly temperature (top) and (bottom) *Pseudomonas* spp. composition based on MAG relative abundance. (B) Summed RPKM values for different *Pseudomonas* spp. (C) Mid-point rooted mash distance tree of 43 *Pseudomonas* MAGs with adjoining model results, including model *R^2^* values for different *Pseudomonas* gs. and coefficients for lagged SXT use. First column is MAG species taxonomic assignment. (D) Example model fits for the best fitting MAGs (*P. mohnii* gs35) and a randomly selected MAG with a significant association with lagged SXT use (gs. 3). Curve and shaded area indicate the predicted seasonality of the MAG, i.e., amplitude and 95% confidence interval. Empty circles: observed values of species abundance; filled circles: model predicted values of MAG abundance. Model fits for all MAGs are available in Supplementary Materials.

Comparisons of **Models 2** and **3** highlighted differences in model fit across species **(Figure 5C, Supplementary** Figure 13). Significant improvement to model fits were observed **(Fig. 5C),** most strongly for MAGs classified to *P. fluorescens* and *P. mohnii* **(Supplementary Data 6-8)**. Our results highlighted a subset of genomes, most prominently *P. mohnii* gs35, for which lagged SXT use was a significant predictor **(Fig. 5D).** The observed trends were again consistent across different species considering **Model 4,** which adjusted for multiple potential explanatory variables. The coefficient for SXT use in *P. mohnii* gs35 from **Model 3** was 1.57 (95% confidence interval: 1.17 – 1.97), implying a ∼1.6 unit increase in *P. mohnii* abundance for one 1 unit increase of SXT use. Effect sizes, including partial Cohen’s *f^2^* and η^2^, were calculated which reaffirmed the strong explanatory effect of lagged SXT usage **(Supplementary Tables S2 and S3).** Species without significant seasonality (amplitude FDR < 0.05) included *P. coleopterorum, P. fluorescens BC,* among others.

Interestingly, no *Pseudomonas* MAGs displayed statistically significant negative correlations with lagged SXT use, suggesting that antibiotic use may have had a promotive effect on some *Pseudomonas* species, but no effect on others **(Fig. 5D, Supplementary Table 4)**. Negative coefficient estimations were observed for some species, e.g., *P. coleopterorum* gs19, however, model performance for this species was very poor (R^2^ < 0.1). Influent flow was hypothesized to be related to *Pseudomonas* abundance due to shear stress on pipe surfaces, however, only 8 MAGs displayed significant associations in **Model 4** with coefficients ranging from 0.06 – 0.1 **(Supplementary Table 5).** Of these 8, 3 were not associated with crAssphage relative abundance, meaning that their profiles were not detectably attributable to fecal input **(Supplementary Table 6)**. This included *P. mohnii* gs35. No MAGs displayed statistically significant associations with day of the week **(Supplementary Data 8).** We also tested the hypothesis that SXT use was associated with a decreasing abundance of *Escherichia coli* (a typical target of SXT and causative agent of urinary tract infections) by modeling the abundance of MAG *E. coli* gs. 45 using **Model 4.** However, while we observed a negative coefficient value for lagged SXT use, it was not statistically significant and the overall model performance was poor, except for a strong association with crAssphage **(Supplementary Data 9).** This may be partly due to an elevated rates of resistance (∼75% susceptible) in the area.

Finally, we investigated whether there was a genomic basis for increased resistance in *Pseudomonas* gs. that had significant associations with lagged SXT use relative to those that did not. Again, only one *P. alcaligenes* MAG was considered to avoid biasing the comparisons. It was noted that genomes with direct associations with lagged SXT use had overall larger genomes (Wilcoxon Rank-sum test: median 5,145 genes vs. 3,341 genes; n = 33; p < 0.01) **(Supplementary** Figure 14). Interestingly, genomes with significant associations displayed an increased rate of carriage of genes encoding efflux pumps within COG category V (defense mechanisms) (**Supplementary** Figure 15), but not efflux-related genes in any other COG category (median 15 genes per genome vs. 10, n = 33; p < 0.01). Similarly, genes encoding COG category F proteins (nucleotide transport and metabolism) were enriched in directly associated gs. **(Supplementary** Figure 16) (145 vs. 94, n=33; p<0.001). These results were not statistically significant when considering gene counts as a proportion of total genes, suggesting that genome size was the underlying driver of statistical significance. However, this does not preclude a model wherein *Pseudomonas* species with larger genome sizes encode more proteins related to environmental nucleotide scavenging as well as an increased assortment of efflux pumps, a key determinant of AR.

### Disentangling ecological and epidemiological signals of AR in sewage

Here we address the role of sewage microbial ecology as a governing factor shaping AR indicators measured in sewage and identify field-scale evidence of antibiotic use-driven selection pressure favoring the growth of certain resistant bacteria in the sewage collection system. Furthermore, we identify a subset of the resistome that appeared to derive from the human gut and which displayed strong associations with antibiotic use. These trends were strongly linked to the putative hosts of ARGs, and multiple co-occurring CAGs of microbes with varying associations with seasonal traits (e.g., temperature) and crAssphage were found that mirrored trends in the resistome. Interestingly, we found that *Enterobacteriaceae-*associated ARGs tended to have direct associations with antibiotic use, while *Pseudomonadaceae* displayed lagged associations with usage **(Figure 3)**. These trends were also, in part, correlated with the environmental stability and/or bioavailability of the antibiotic, supporting the hypothesis that antibiotic use selects for resistance, both within the community and in the sewer environment. By leveraging joint modeling of physical-chemical sewage parameters, seasonality, and antibiotic use, we identify measured relationships between community antibiotic use and AR indicators in sewage. These analyses were made possible via a unique dataset including sewershed-specific antibiotic prescription rates from outpatient settings comprising the majority of human antibiotic consumption in the community.

Unlike some previous studies (11, 13, 14), we observed a strong relationship between AR measures in sewage and antibiotic use. This difference is likely attributable to our longitudinal study design, selection of continuous response variable (individual, cell-normalized ARG relative abundance), and the availability of comprehensive antibiotic use data. We suggest that individual ARGs, as opposed to drug class abundances, capture more variability driven by within-patient selection during antibiotic therapy. This finding is important for current efforts to design and implement effective WBS for AR in situations where metagenomics is not feasible and specific gene targets are pre-specified. In our study, ARGs associated with fecal-associated microbiota (e.g., *Enterobacteriaceae*) were well-supported as markers of community resistance. In contrast, genes with broad host ranges (e.g., *mphA, sul2*, and “older” ESBLs) are less likely to reflect ongoing shifts in community AR and may have limited relevance as markers for community resistance. In sum, the results of the present study support using clinical data to identify correlates of sewage-associated AR to aid in the identification of suitable indicators for WBS.

Considering the value of WBS for AR, it is worth highlighting the extensive collection of MAGs recovered in the present study and elsewhere. In this context, MAGs—and the metagenomic assemblies—are essentially hypotheses regarding the associations of specific resistance phenotypes with genotypes. Towards the future, bioinformatic method development aimed at improving the capacity to link these genotypes to phenotypes could amplify the public health value of WBs by providing a new means to inform regional clinical practice in light of circulating resistant strains.

## METHODS

### Field sampling and sequencing

Twice-weekly sampling of influent wastewater was conducted at a small local WWTP in southwestern Virginia serving approximately 20,000 people for a period of one year. Mixed ester cellulose 0.22 µm filters were washed with autoclaved nanopure water before filtering influent and effluent wastewater, until clogging of the filter occurred. A lab blank was also included for each sampling point which received only autoclaved nanopure water. To capture potential contaminants incurred during sample preparation, DNA extraction, and sequencing, we added 37 µL of the Zymo Mock Microbial Community (DS6700, Zymo Research, Irvine, CA) whole cell spike into the lab blank filter tube prior to extraction. For sequencing, a representative sample of about 10 lab blanks were combined and submitted for sequencing in parallel for each flow cell.

DNA was extracted using the FastDNA SPIN-kit for soil (MP Biomedicals, Solon, OH) with modifications. We increased the first centrifugation step by 10 minutes to increase separation of filter fragments from soluble supernatant. Final elution was conducted in molecular grade water. DNA was quantified using a QuBit 4 fluorometer (Thermo Fischer Scientific, Waltham MA) with the high sensitivity dsDNA detection assay kit from Thermo Life Sciences (Q33120, Thermo Fisher, Waltham MA). We confirmed the integrity of the extracted DNA for a subset of samples over the length of the sampling period by gel electrophoresis of purified genomic DNA. Samples were then submitted to Duke for Tagmentation library preparation and sequencing for 150 bp paired end sequencing on an Illumina NovaSeq6000 (Illumina, San Diego, CA) with S4 chemistry flow cells.

### Preprocessing and data analysis

The preprocessing steps taken here were based on the JGI metagenome workflow^14,15^ and includes reference based decontamination. Quality control of raw reads was performed using fastp, –-trim_poly_g, –-detect_adapter_for_pe, –-trim_poly_x, –-average_qual 10) and then decontamination of the reads was performed using bbduk (with *k*-mer size of 51) using the masked human, rat and cow genomes associated with the bbtools suite. Assembly of the Illumina metagenomes was performed using megahit (42) (--meta-sensitive). ARGs in the short reads were quantified using both CARD (43) v.3.1.4 and deepARG-db (44) (diamond (45) blastx –e 1e-10 –-id 80 –k 1) while 16s rRNA gene copies were calculated by aligning the short reads to GreenGenes (46) v.13.5 using minimap2 (47) (-x sr; script: run_minimap2.sh). Open reading frames were predicted for each sample’s contigs using prodigal (-p meta). For ARGs, 16s rRNA normalized ARG counts, reads per kilobasepair million (RPKM), fragments per kilobasepair million (FPKM), and *rpoB* normalized data were calculated using rel_abund.py. Taxonomic profiles of the short reads were calculated using kraken2 with both the default (NCBI-derived) taxonomy database as well as GTDB (48, 49) (v.202) with default settings. We clustered the CARD v.3.1.4 database using mmseqs2 (50) (-c 0.8 –-id 90 –-cov-mode 1). We also manually allocated major drug classes to ARGs based on the gene family and resistance mechanism **(Supplementary Methods 1).**

### Assembly and binning

We produced a catalogue of sewage-associated ARGs from the metagenomic data. We attempted to use metaSPAdes, which includes scaffolding steps, but this proved to be computationally infeasible because of the size of the data, despite access to more than 900 Gb of RAM. Thus, we implemented a reassembly strategy that (1) extracted reads from samples that aligned to the megahit contigs bearing ARGs using bowtie2 and samtools; (2) harmonized extracted paired-end reads using fastq-pair (51); and (3) re-assembled the short reads with the total catalogue of nanopore long reads using hybridSPAdes. The final assembly catalogue was dereplicated at 99% identity using mmseqs2 (52) and taxonomically annotated using mmseqs taxonomy (53).

Using the original megahit assemblies, contigs with length greater 1500 bp were binned using metabat2 using all samples with mash distance of less than 0.028 relative to the origin read sample set **(Supplementary Methods 2).** Resulting bins were dereplicated using dRep (54) v.3.4.0 (-comp 50 –con 25 –-S_algorithm fastANI –-multiround_primary_clustering –ms 10000 –pa 0.9 –sa 0.95 –nc 0.30 –cm larger). Taxonomy was assigned to the dereplicated bins using GTDB-Tk v.2.3.2(48, 49) with the MASH(55) reference sketch database v2.3 and the GTDB-Tk reference data version v.r207.

### Coabundance group prediction

CAGs for MAGs and ARGs were predicted using the elbow method to identify best fit number of clusters as described before (56). Briefly, correlations were calculated in R using cor(method=”spearman”) and converted to a distance matrix using vegdist(method=”euclidean”) from vegan v2.6-4 (57), followed by hclust(method=”complete”). Final clusters were picked based on a visualization of the Dunn index.

### Outpatient antibiotic prescriptions

Antibiotic prescriptions at local outpatient clinics collected by electronic filing were summarized to provide a sewershed-specific dataset from patient self-reported home address. Mean daily prescriptions per 10,000 visits were calculated for the sewershed associated with the WWTP, and the number of visits summed across all clinics within the zip-codes of the sewershed.

### Antibiotic prescription modeling

We tested for seasonality of 13 antibiotic sub-classes: aminopenicillins/aminopenicillins with beta-lactamase inhibitors; antibacterial folate antagonist (other combinations); antiprotozoal-antibacterial 1^st^ generation 2-methyl-5-nitroimidazole; cephalosporin antibiotics (partitioned by 1^st^, 2^nd^, and 3^rd^ generation); fluoroquinolones; lincosamide, macrolides, tetracyclines, penicillins, and nitrofurans.

Outpatient antibiotic usage was fit to a model for seasonality as described in Sun *et al* (30) **(Model 1).** All models were fit using nonlinear least square regression using nls from the stats package (v. 3.6.2) in R. Code used in modeling is available: https://github.com/clb21565/WBS_use_resistance_modeling.

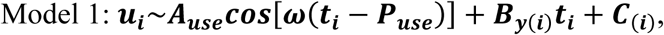

where *u_i_* is the mean daily prescriptions per 10,000 visits in calendar month *t_i_*; *A_use_* is the amplitude of seasonality for usage; *P_use_* is the phase of use; 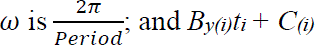 is a linear component for the month *t_i_*.

### MAG relative abundance modeling

The relative abundance of specific *Pseudomonas* spp. was modeled considering multiple factors, including antibiotic use, day of week associated with sampling point, and level of crAssphage using

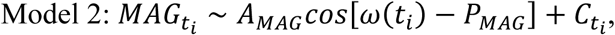

where *MAG_ti_* is the relative abundance of a particular MAG at month *t_i_* (in RPKM)*, A_mag_* is the amplitude of seasonality for *MAG_i_*; *P_mag_* is phase, and 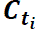 is an intercept term for baseline variability in the study. We fit additional models to test the effect of antibiotic usage directly on *Pseudomonas* spp. abundance while controlling for seasonality and various extrinsic factors.

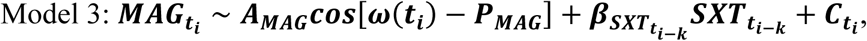

where 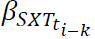 is the coefficient for antibiotic usage, 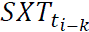 is the antibiotic usage at time 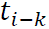 (in mean daily prescriptions per 10,000 visits, where *k* is the lag value in months); and 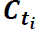 is an intercept term for baseline variability in the study.

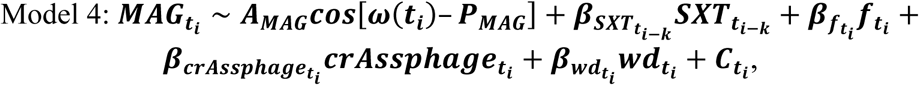

where 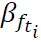 and 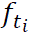 are the coefficient and value for influent flow, respectively; 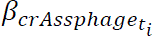 and 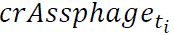 are the coefficient and value for crAssphage abundance, respectively; and 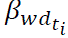 and 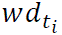 are the coefficient and a binary coded variable for Monday (0) or Friday (1), respectively. Influent flow was included as it is related to the pipe shear stress exerted on biofilms, likely impacting the release of biofilm-associated microbiota in sewers (9, 10).

## Ethical Approval

The study was deemed exempt by the Carilion Clinic Institutional Review Board (IRB-22-1775) as it is not categorized as human subjects research as it included only de-identifiable prescription data.

## Acknowledgements

The authors acknowledge Daphne Sun for her suggestions for modeling analyses. The research presented was not performed or funded by EPA and was not subject to EPA’s quality system requirements. The views expressed in this article are those of the author(s) and do not necessarily represent the views or the policies of the U.S. Environmental Protection Agency.

## Data Availability

All sequencing data are available via BioProject PRJNA1083020.

