## Supplementary Information for "Metagenomics disentangles epidemiological and microbial ecological associations between community antibiotic use and antibiotic resistance indicators measured in sewage"

### Supplementary Materials

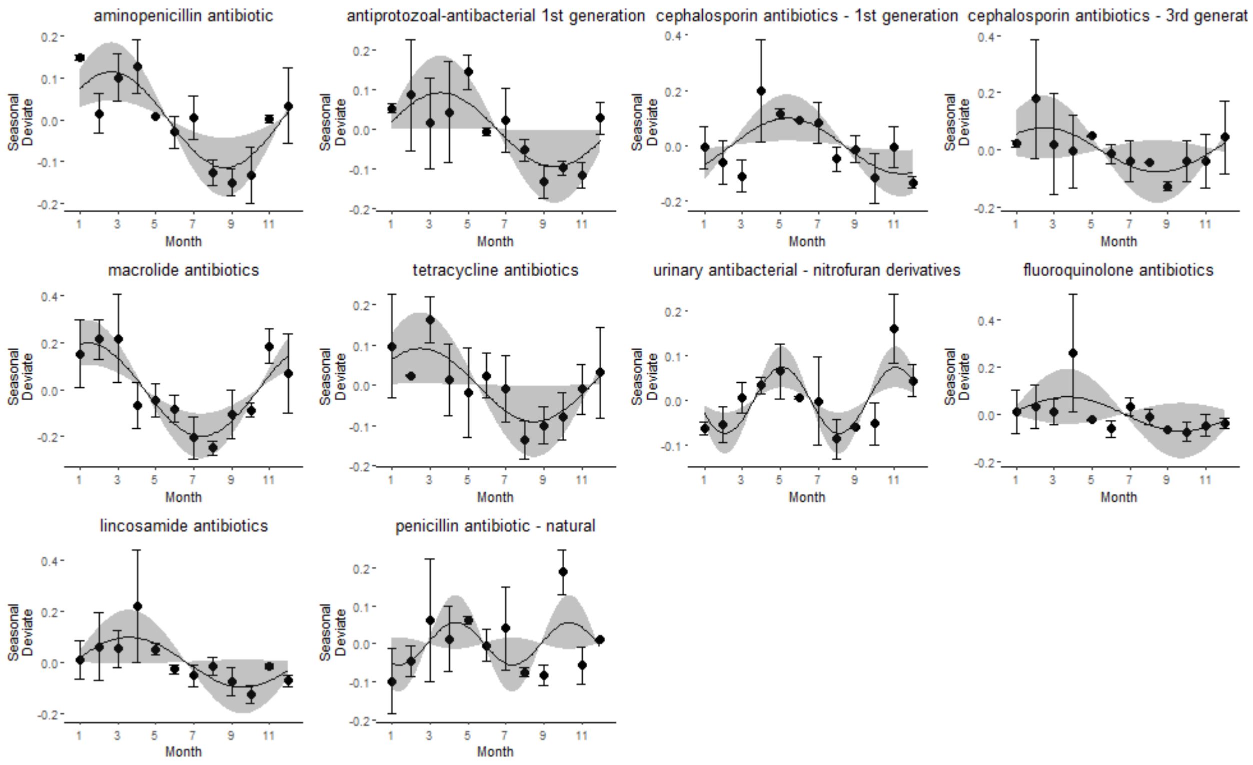


Supplementary Figure 1. Drug classes with statistically significant seasonality using data from 2020 and 2021. Points are seasonal deviates of use representing deviations from the yearly average. Error bars are standard error of the mean (SEM) across all years. Curve represents the model fitted values for each month and the shaded areas indicate 95% confidence intervals.

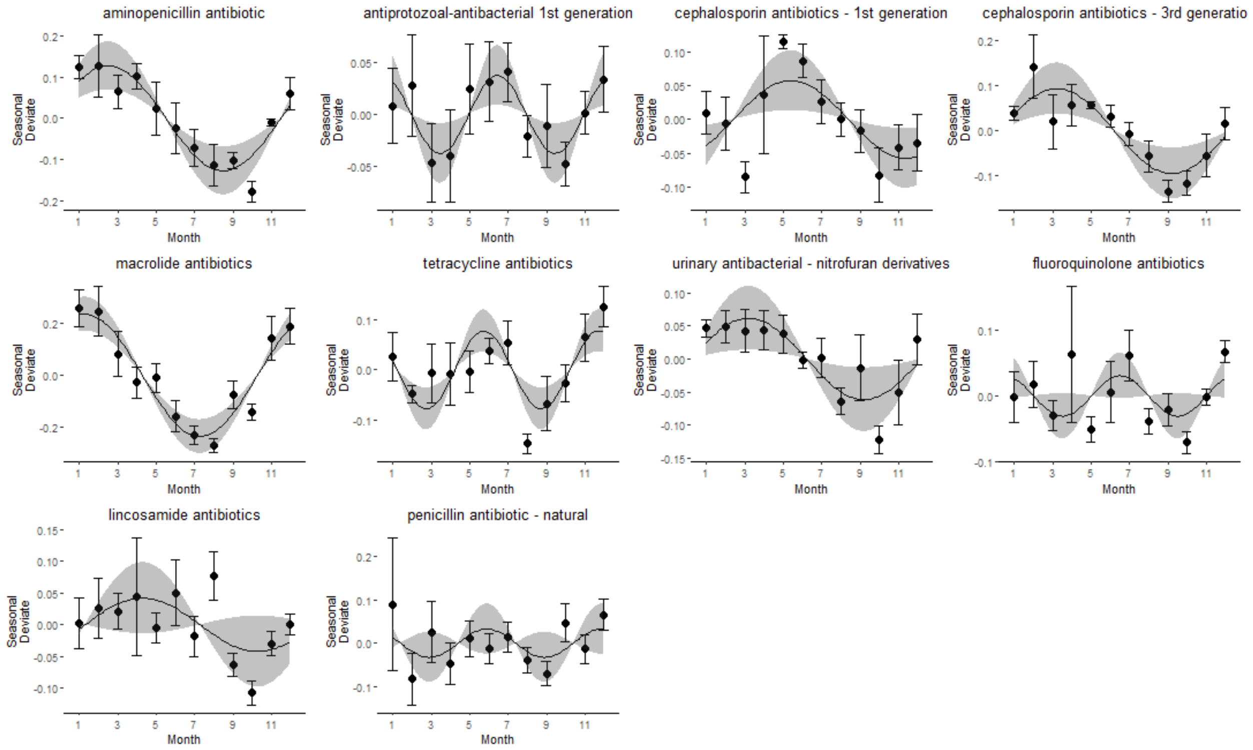


Supplementary Figure 2. Drug classes with statistically significant seasonality using data from 2017-2022. Points are seasonal deviates of use representing deviations from the yearly average. Error bars are standard error of the mean (SEM) across all years. Curve represents the model fitted values for each month and the shaded areas indicate 95% confidence intervals.

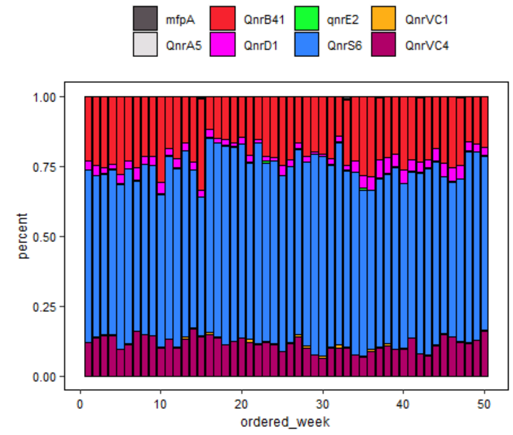
mfpA
QnrB41
qnrE2
QnrVC1
QnrA5
QnrD1
QnrS6
QnrVC4
percent
ordered_week

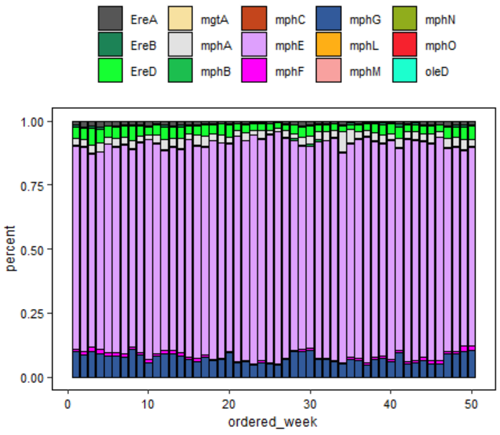
EreA
mgtA
mphC
mphG
mphN
EreB
mphA
mphE
mphL
mphO
EreD
mphB
mphF
mphM
oleD
percent
ordered_week

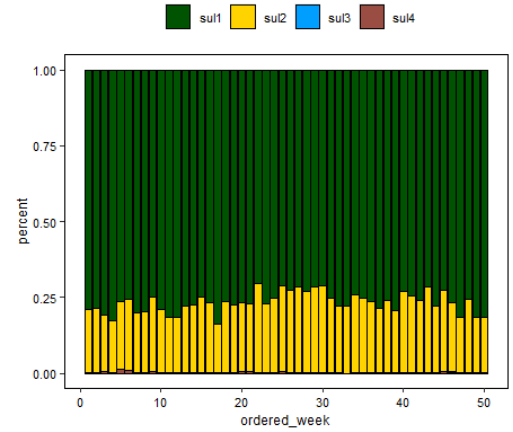
sul1
sul2
sul3
sul4
percent
ordered_week

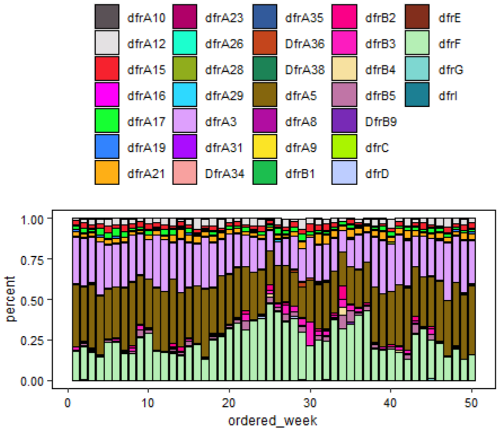
dfrA10
dfrA23
dfrA35
dfrB2
dfrE
dfrA12
dfrA26
DfrA36
dfrB3
dfrF
dfrA15
dfrA28
DfrA38
dfrB4
dfrG
dfrA16
dfrA29
dfrA5
dfrB5
dfrI
dfrA17
dfrA3
dfrA8
DfrB9
dfrA19
dfrA31
dfrA9
dfrC
dfrA21
DfrA34
dfrB1
dfrD
percent
ordered_week

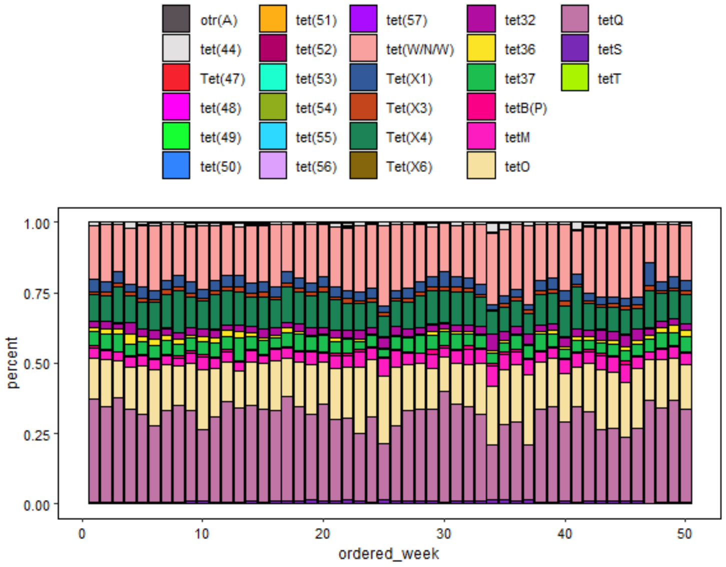
otr(A)
tet(51)
tet(57)
tet32
tetQ
tet(44)
tet(52)
tet(W/N/W)
tet36
tetS
Tet(47)
tet(53)
Tet(X1)
tet37
tetT
tet(48)
tet(54)
Tet(X3)
tetB(P)
tet(49)
tet(55)
Tet(X4)
tetM
tet(50)
tet(56)
Tet(X6)
tetO
percent
ordered_week

Supplementary Figure 3. Drug class abundance is dominated by a handful of abundant gene families. The y-axis reflects the proportion of abundance associated with a given ARG cluster (delineated by color). The x-axis refers to the week of sampling. Drug classes (from left to right) are quinolone, macrolide, (second row) sulfonamide, diaminopyrimidine, and (last panel) tetracyclines.

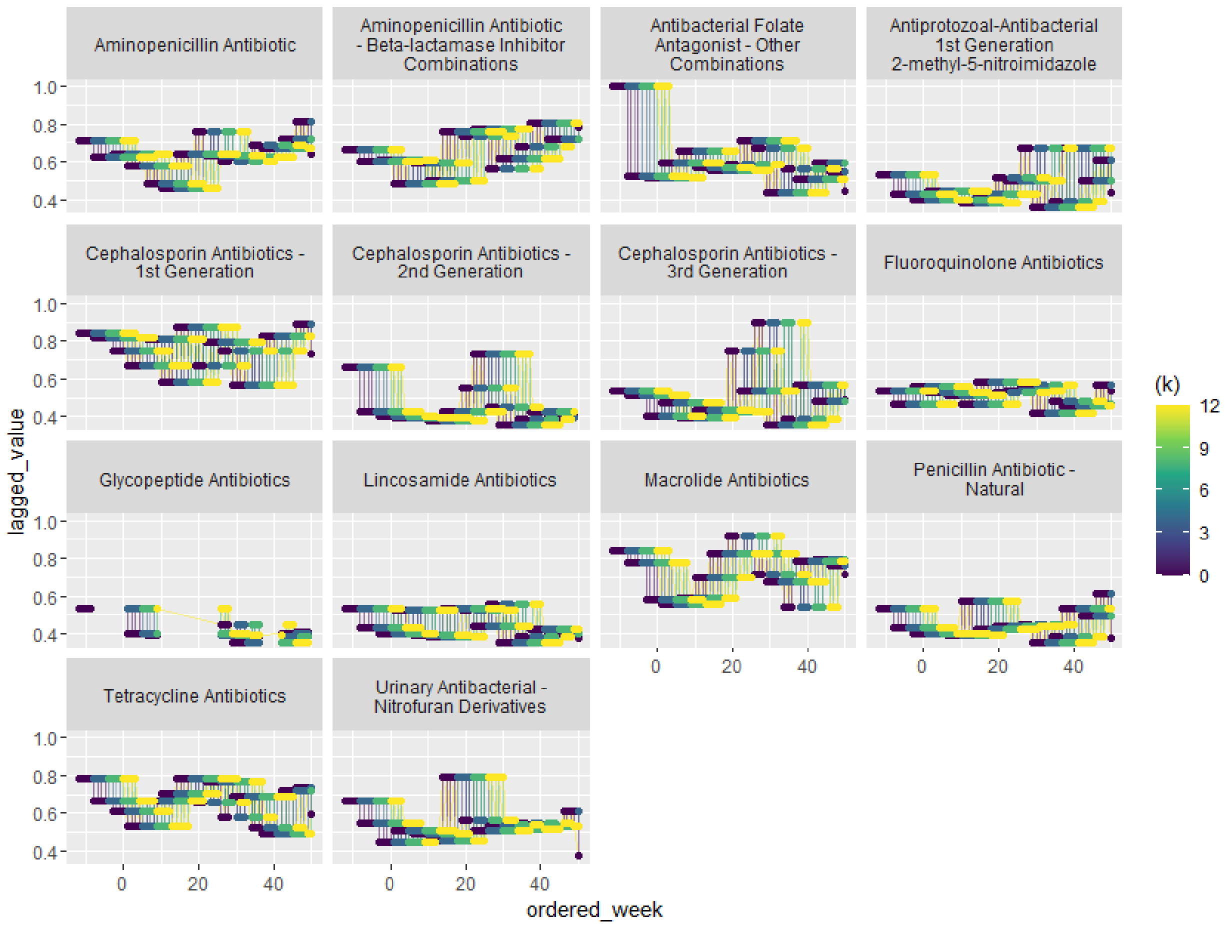


Supplementary Figure 4. Lagged usage data including up to 3 months prior to the start of sampling. The heat (*k*) refers to the number of weeks the data were lagged by.

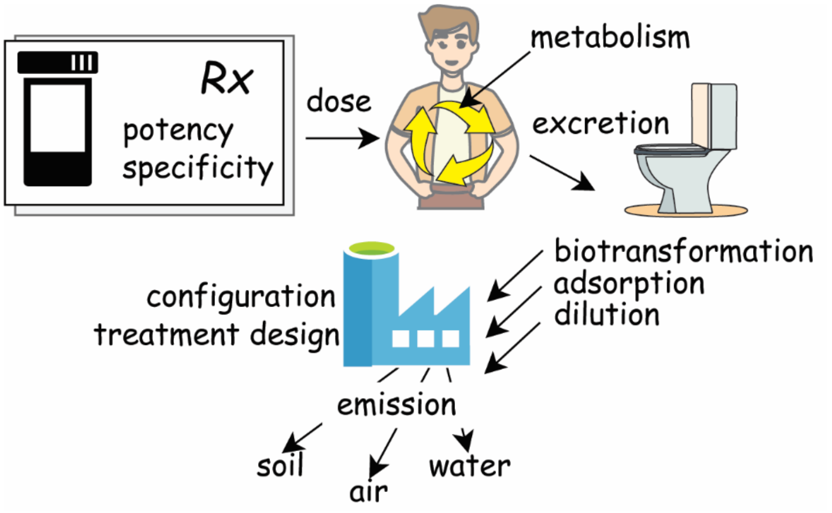


Supplementary Figure 5. Life Cycle of antibiotics illustrates important factors contributing to their environmental concentration and bioactivity.

Supplementary Table 1. Hypothetical factors underlying antibiotic-driven selection for antibiotic resistance along the life-cycle of antibiotics.

**Table 1. Hypothetical factors underlying antibiotic-driven selection for antibiotic resistance along the life-cycle of antibiotics.**

| Factors | Notes |
|---|---|
| Potency of the drug; prescribed dosage; length of therapy; pharmacological specificity | Therapeutic treatment with antibiotics requires sufficient dosage to provide its inhibitory effect<br>Environmental concentrations of antibiotics are expected to always be far below therapeutic doses in municipal wastewater<br>Potency can mean multiple things; could mean it has a low MIC value; could also mean it is selective against a broad range of potential targets |
| Amount of drug metabolized/excreted; amount of drug persistent in active/bioavailable state | Antibiotics display different stabilities in the environment and undergo transformation both within humans as well as the environment<br>Transformation and excretion kinetics of different antibiotics (in addition to which excreta they will be associated with) vary<br>The antimicrobial activity of a given antibiotic is impacted by any metabolism or transformation |
| Adsorptive or other physicochemical properties | In addition to environmental stability, different antibiotics display different physicochemical properties that impact their fate and transport during conveyance<br>Excreted drug must be able to reach the susceptible microbes in bioavailable form<br>Different organisms partition more/less strongly to biofilm communities in sewer and suspended particles<br>Adsorption to biofilms could create a lagging impact for the influent signal<br>Adsorption to biofilms could have a net positive or net negative impact on antibiotic resistance by either (1) removing antibiotic contamination from aqueous fraction and/or (2) promoting resistance in sewer biofilms |

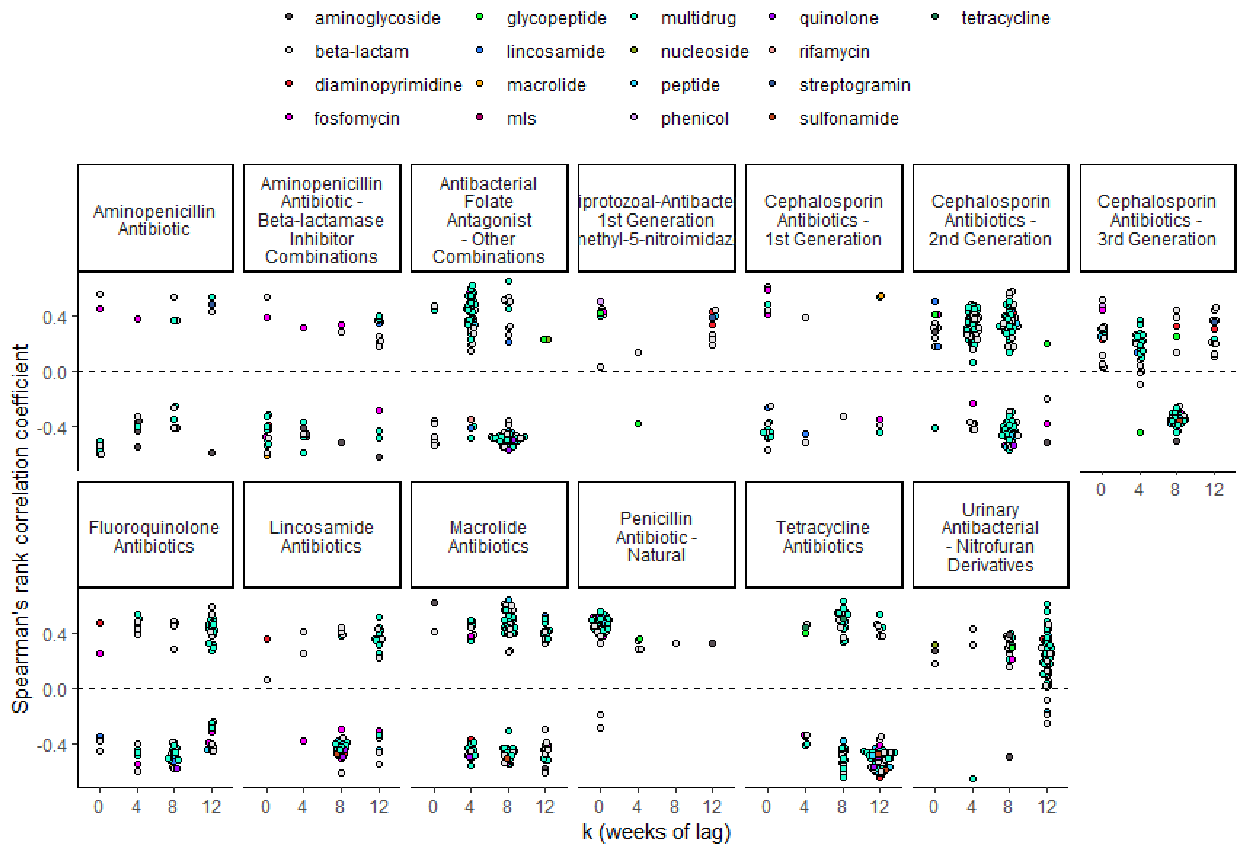


Supplementary Figure 6. Antibiotic usage displayed heterogeneous relationships with ARGs. Results of multiple comparisons of ARG abundances with antibiotic usage considering 0, 4, 8, or 12 weeks of lag.

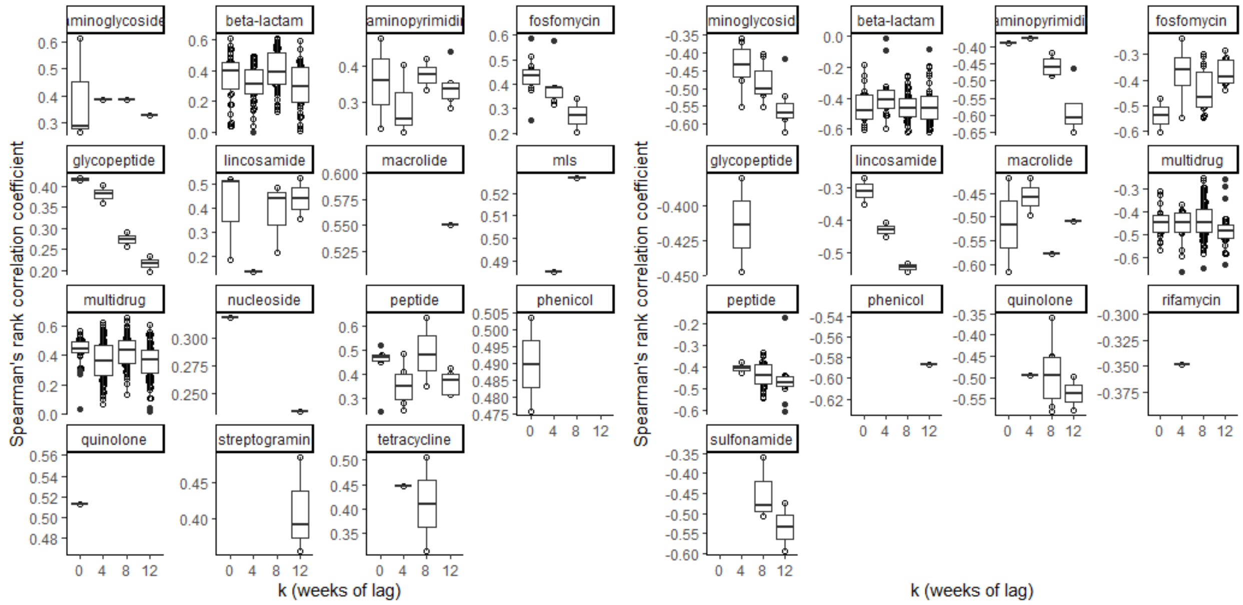

Supplementary Figure 7. Correlation between antibiotic usage and ARGs of different drug classes. Left panel are genes with statistically significant positive coefficients. Right side depicts genes with negative coefficients.

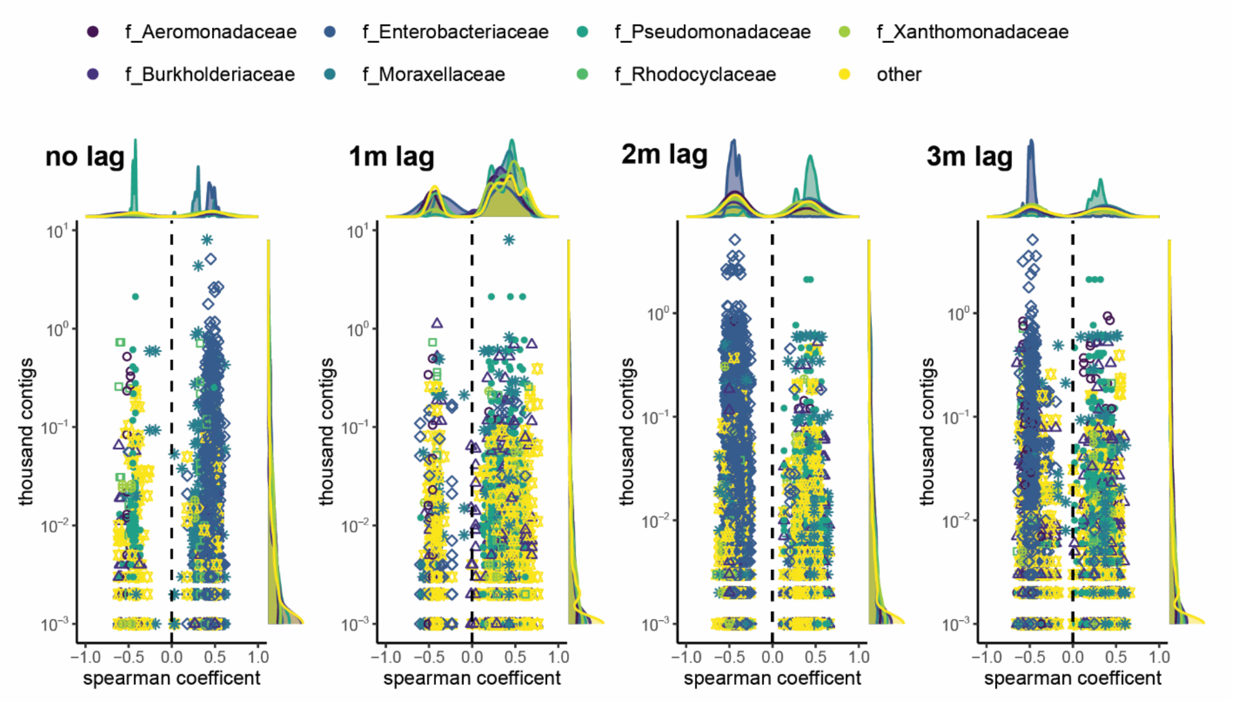


Supplementary Figure 8. Pattern of correlation with usage is linked to host of ARGs. Hosts of ARGs were predicted by annotating contigs derived from metagenomic assemblies (see Supplementary Methods). Taxonomic information regarding the top 7 most frequent ARG hosts (at the family level) is overlaid onto correlation coefficients. Each dot represents one ARG-taxa pairing and the y-axis relays the number of contigs that had that ARG and were found to be labeled with the respective taxonomic assignment.

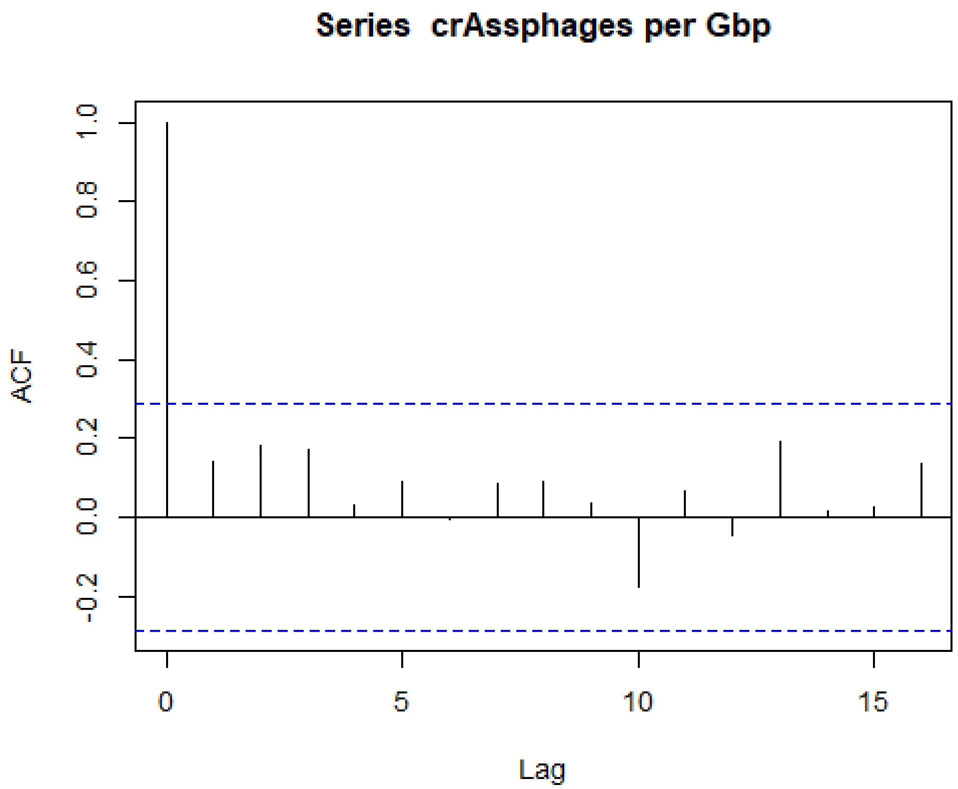


Supplementary Figure 9. crAssphage ACF plot shows no signs of autocorrelation.

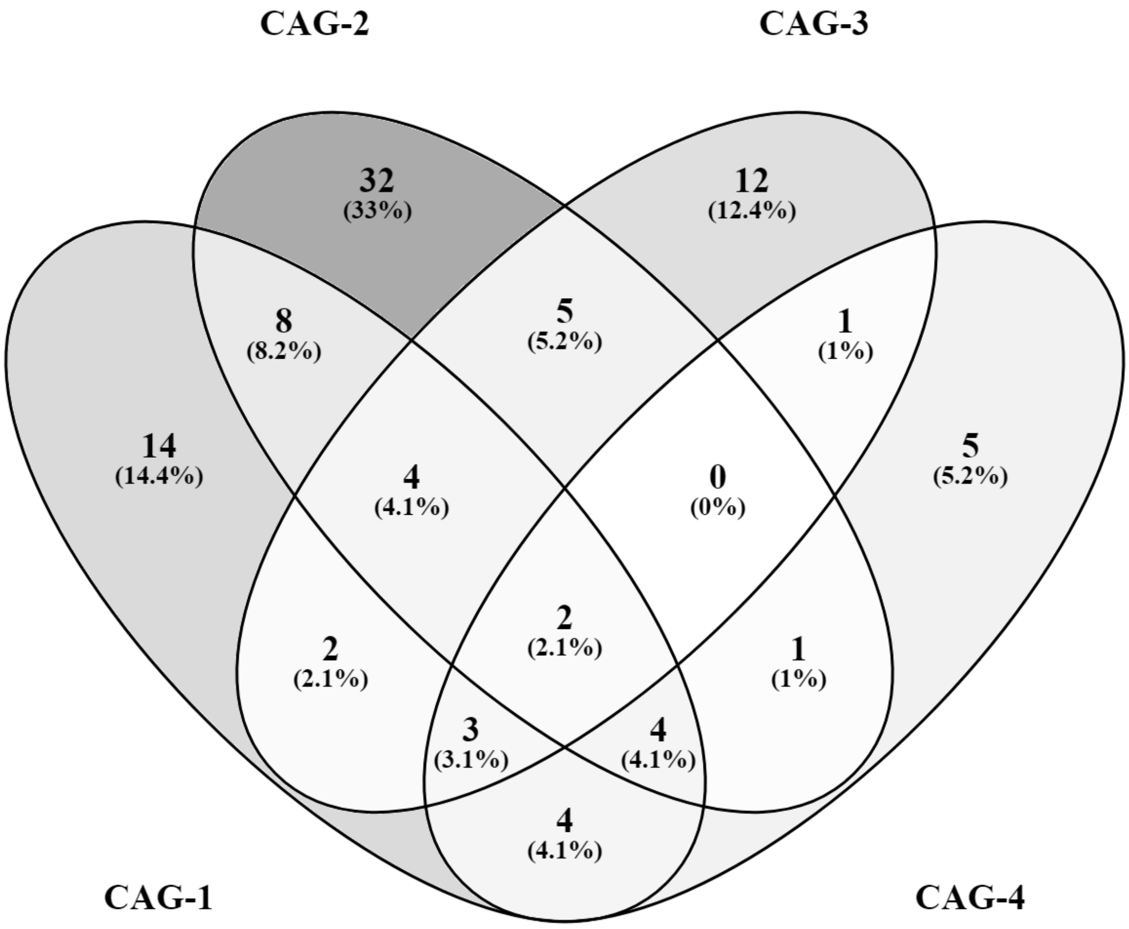


Supplementary Figure 10. Overlap of co-abundance group genera.

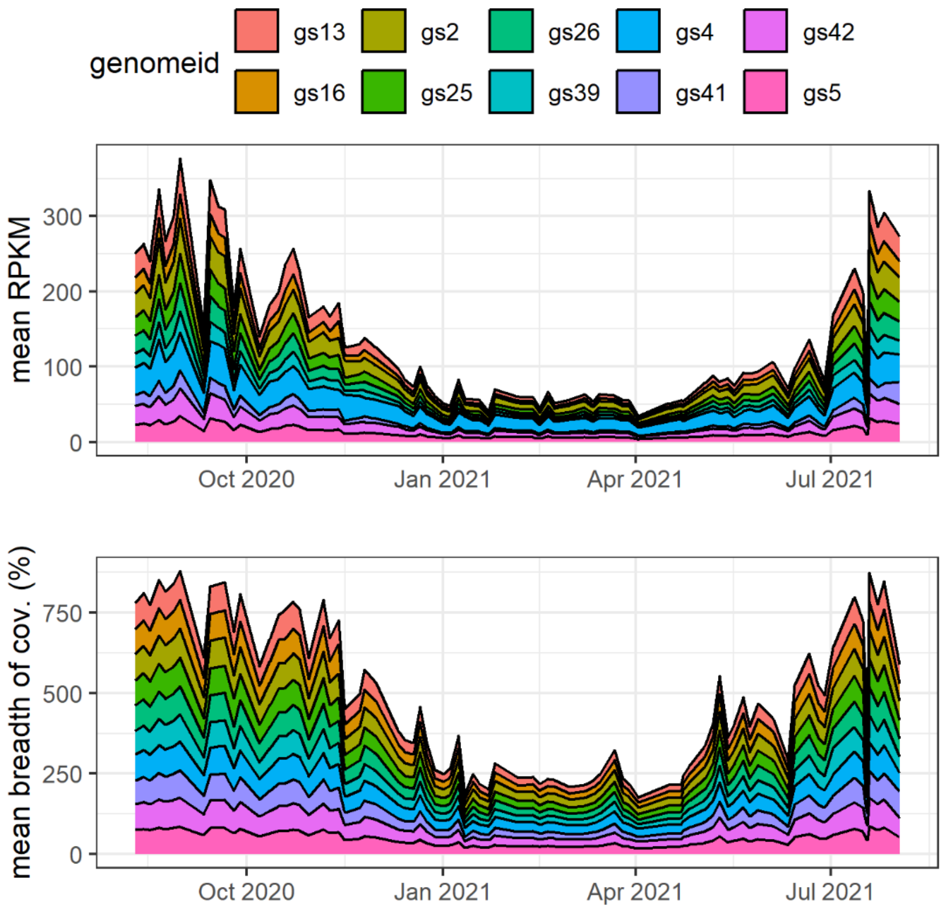


Supplementary Figure 11. Relative abundances and mean breadth of coverages for *Pseudomonas alcaligenes* genomospecies (n = 10).

Supplementary Table 2. SXT coefficient values from models where SXT was a statistically significant model parameter

| Table S2. **SXT coefficient values from models where SXT was a statistically significant model parameter.** | | | |
|---|---|---|---|
| | | Slope (95% CI) | |
| Genome | Species | Model 3 | Model 4 |
| gs35 | *P. E mohnii* | 1.57 (1.17,1.97) | 1.61 (1.2,2.03) |
| gs30 | *P. E sp900187645* | 1.52 (1.11,1.93) | 1.51 (1.09,1.94) |
| gs31 | | 1.5 (1.09,1.92) | 1.3 (0.89,1.72) |
| gs7 | *P. E fluorescens BJ* | 1.47 (1.09,1.84) | 1.42 (1.02,1.82) |
| gs36 | | 1.46 (1.05,1.86) | 1.26 (0.85,1.67) |
| gs33 | *P. E mohnii* | 1.39 (1.01,1.76) | 1.36 (0.96,1.76) |
| gs14 | | 1.33 (0.94,1.72) | 1.39 (0.99,1.79) |
| gs34 | *P. E mohnii* | 1.27 (0.83,1.71) | 1.1 (0.65,1.55) |

| | | | |
|---|---|---|---|
| gs29 | | 1.24 (0.9,1.59) | 1.25 (0.87,1.64) |
| gs18 | *P. E fluorescens BC* | 1.24 (0.69,1.78) | 1.08 (0.55,1.61) |
| gs28 | *P. E fluorescens BA* | 1.21 (0.83,1.6) | 1.32 (0.88,1.76) |
| gs15 | | 1.05 (0.62,1.48) | 0.78 (0.31,1.24) |
| gs3 | | 1.05 (0.62,1.48) | 0.79 (0.33,1.24) |
| gs8 | | 1.05 (0.62,1.48) | 0.79 (0.34,1.24) |
| gs12 | | 1.03 (0.6,1.46) | 0.77 (0.32,1.22) |
| gs24 | *P. E sp002874965* | 0.92 (0.5,1.34) | 1.09 (0.62,1.57) |

Supplementary Table 3. Effect sizes for models 2-4 for *P. E mohnii* gs35.

| Table S3. Effect sizes for models 2-4 for *P. E mohnii* gs35. | | | | | | | | |
|---|---|---|---|---|---|---|---|---|
| Parameter | model | genomeid | Cohen's f partial [95% CI] | | | Eta sq partial [95% CI] | | |
| | | | Cohens_f_partial | CI_low | CI_high | Eta2_partial | CI_low | CI_high |
| amplitude | Model 2 | gs35 | 0.99 | 0.72 | 1.25 | 0.49 | 0.34 | 0.61 |
| phase | Model 2 | gs35 | 0.45 | 0.22 | 0.68 | 0.17 | 0.05 | 0.32 |
| intercept | Model 2 | gs35 | 1.10 | 0.82 | 1.37 | 0.55 | 0.40 | 0.65 |
| amplitude | Model 3 | gs35 | 1.30 | 1.00 | 1.59 | 0.63 | 0.50 | 0.72 |
| phase | Model 3 | gs35 | 2.73 | 2.25 | 3.20 | 0.88 | 0.83 | 0.91 |
| B_SXT | Model 3 | gs35 | 0.87 | 0.61 | 1.13 | 0.43 | 0.27 | 0.56 |
| intercept | Model 3 | gs35 | 0.71 | 0.46 | 0.95 | 0.33 | 0.17 | 0.48 |
| amplitude | Model 4 | gs35 | 1.37 | 1.02 | 1.71 | 0.65 | 0.51 | 0.75 |
| phase | Model 4 | gs35 | 2.68 | 2.14 | 3.21 | 0.88 | 0.82 | 0.91 |
| B_SXT | Model 4 | gs35 | 0.97 | 0.66 | 1.27 | 0.48 | 0.31 | 0.62 |
| B_Crass | Model 4 | gs35 | 0.33 | 0.07 | 0.59 | 0.10 | 0.00 | 0.26 |
| B_IF | Model 4 | gs35 | 0.34 | 0.08 | 0.60 | 0.10 | 0.01 | 0.26 |
| DOW | Model 4 | gs35 | 0.14 | 0.00 | 0.40 | 0.02 | 0.00 | 0.14 |
| intercept | Model 4 | gs35 | 0.95 | 0.65 | 1.25 | 0.48 | 0.30 | 0.61 |

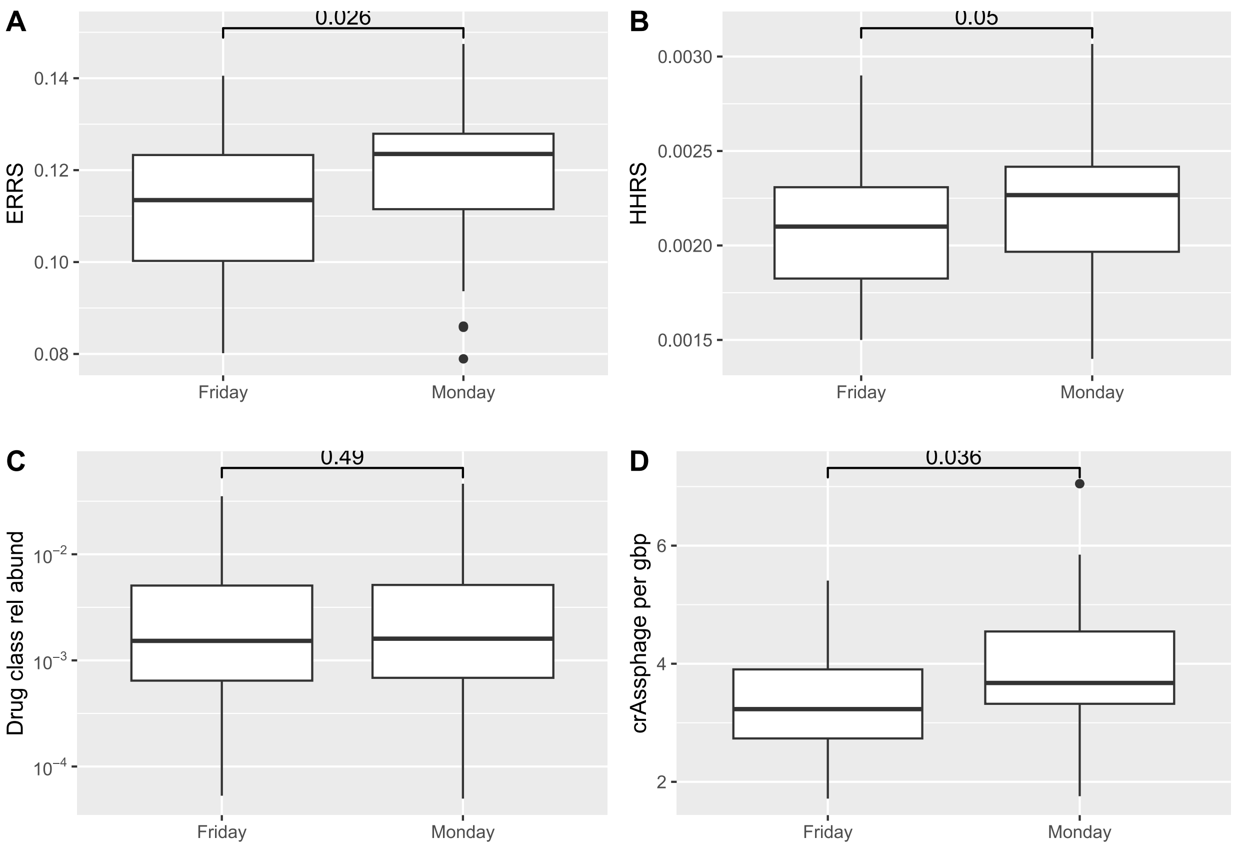


Supplementary Figure 12. Differences in samples collected on Monday vs. Friday. (A) Ecological resistome risk score (ERSS) from MetaCompare2. (B) Human-health resistome risk score (HHRRS). (C) Drug class relative abundance by day of week (*rpoB* normalized). (D) crAssphage relative abundance by day of week.

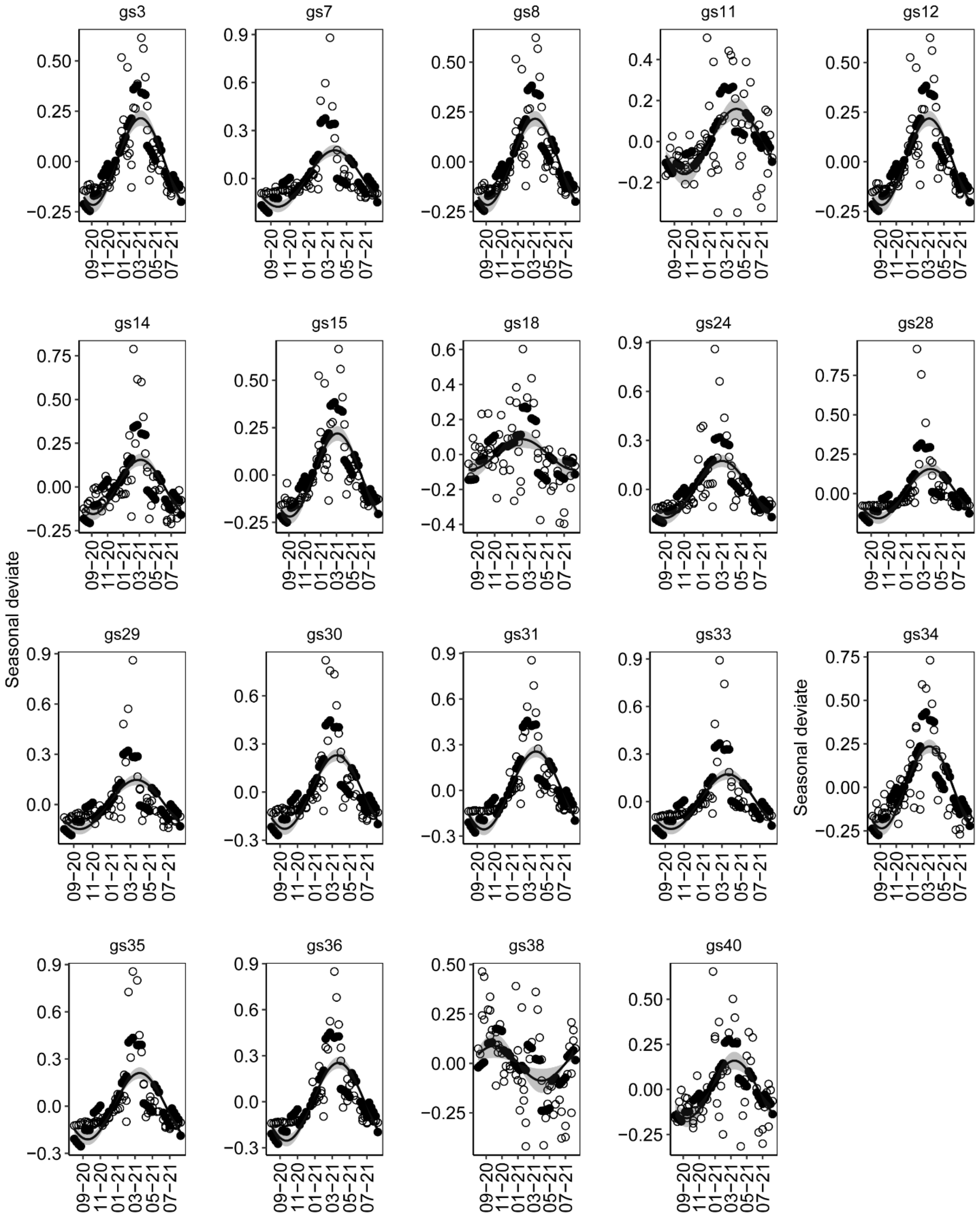
gs3
gs7
gs8
gs11
gs12
gs14
gs15
gs18
gs24
gs28
gs29
gs30
gs31
gs33
gs34
gs35
gs36
gs38
gs40
Seasonal deviate
Seasonal deviate
09−20
11−20
01−21
03−21
05−21
07−21

Supplementary Figure 13. Data fit to model 3. Curve and shaded area indicate the predicted seasonality of the MAG, i.e., amplitude and 95% confidence interval. Empty circles: observed values of species abundance; filled circles: model predicted values of MAG abundance

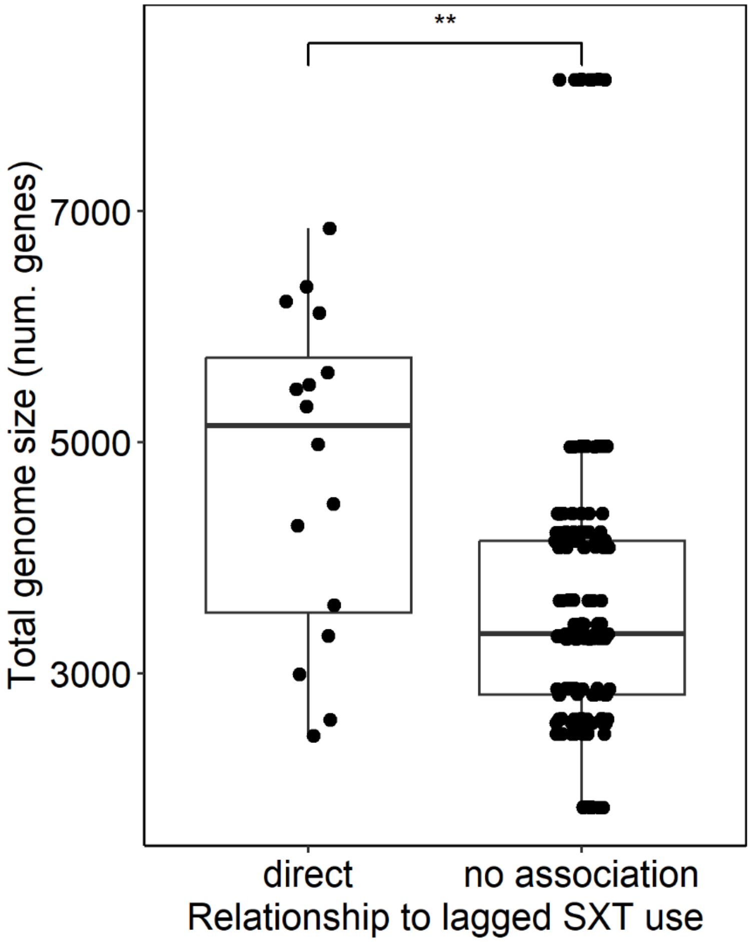


Supplementary Figure 14. Total genome size (in number of genes) of *Pseudomonas* gs. partitioned by their relationship with lagged SXT usage.

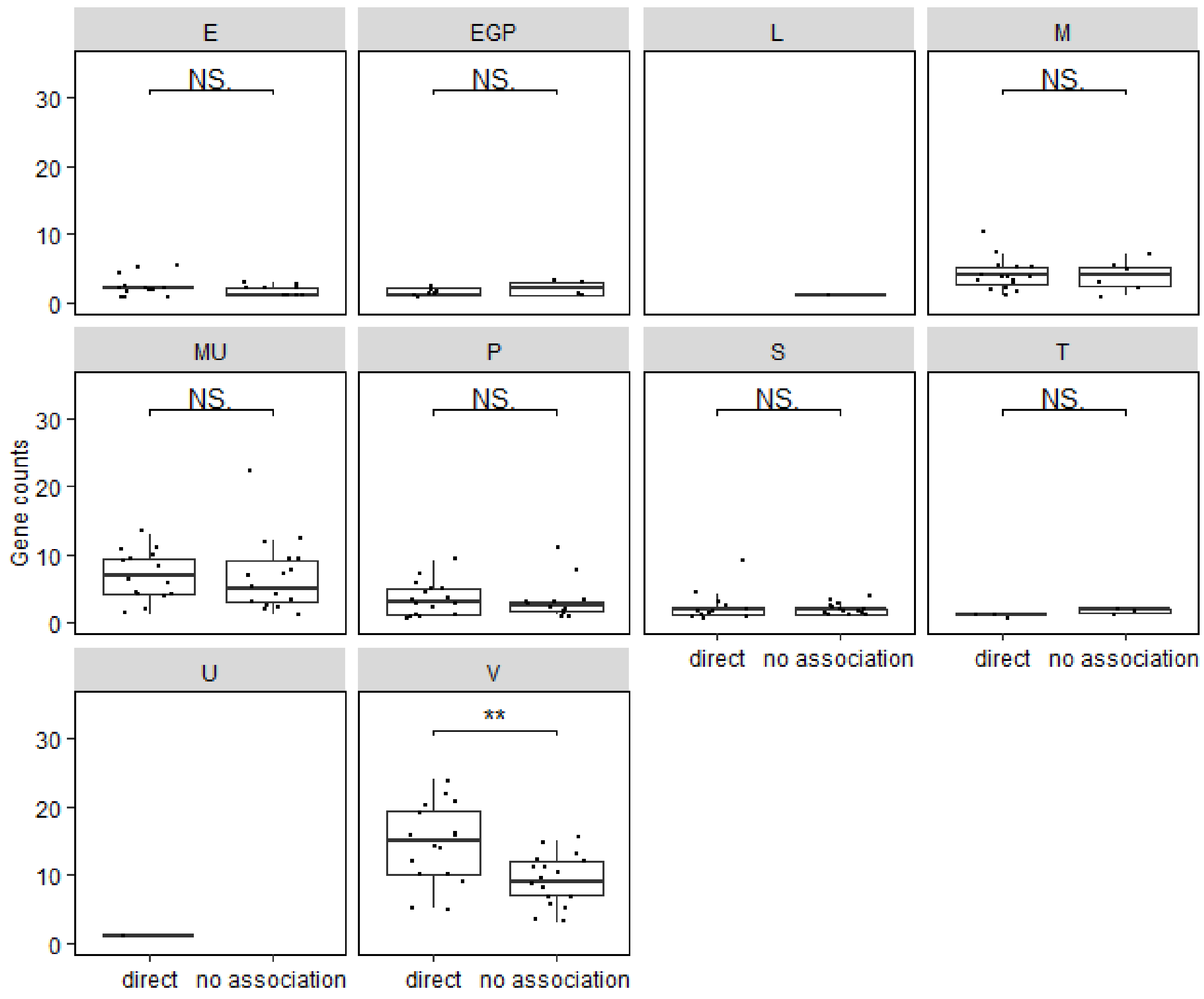


Supplementary Figure 15. Frequencies of efflux related genes in *Pseudomonas* gs. partitioned by their relationship with lagged SXT usage and COG category.

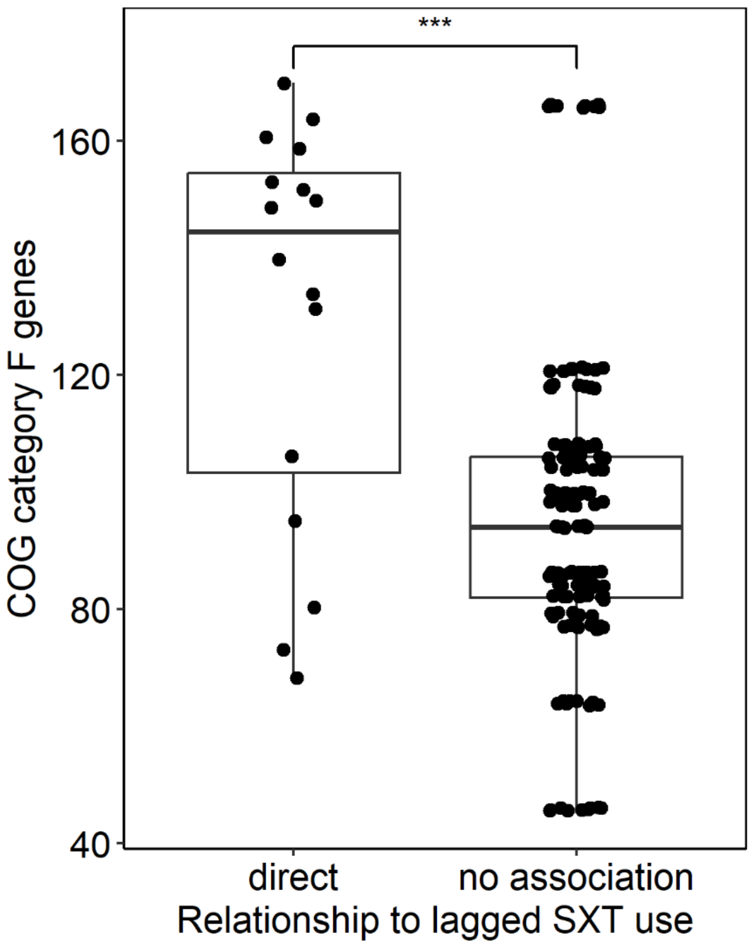


Supplementary Figure 16. Frequencies of COG category F (nucleotide transport and metabolism) genes in *Pseudomonas* gs. partitioned by their relationship with lagged SXT usage.
